# Wastewater and Environmental Surveillance in Cities with Sewered and Non-sewered Sanitation: Evidence from Kampala, Uganda

**DOI:** 10.64898/2026.08.27.26360080

**Authors:** Seju Kang, Annet Kagene, Gava Job Ssazi Pius, Jude Zziwa Byansi, Gloria Mirembe, Flavia Zabali Musisi, Charles B. Niwagaba, Karin Gallandat, Timothy R. Julian, Linda Strande

**Author notes:** Co-corresponding authors and Postal address: Überlandstrasse 133, CH - 8600 Dübendorf.

## Abstract

**Background:** Wastewater and environmental surveillance (WES) enables community-level monitoring of infectious diseases. Most progress has focused on sewer-based surveillance, yet nearly half of the global population relies on non-sewered sanitation. In non-sewered settings, urban drainage channels have been used for poliovirus environmental surveillance, but their potential for a multi-pathogen WES with spatially defined catchments, and comparability to sewer-based surveillance, remains under-explored.

**Methods:** Ten drainage channel sampling points with delineated micro-catchments (0·95-3·83 km^2^ with 12,885-44,146 people) were selected within Kampala. A total of 255 drainage channel and 54 wastewater treatment plant (WWTP) influent samples were collected during two campaigns in March and September-October 2025. A multi-target panel was quantified by digital PCR, including enteric viruses (Norovirus GI and GII, Rotavirus), respiratory viruses (SARS-CoV-2, Influenza A and B viruses, Respiratory Syncytial Virus (RSV)), non-O1/O139 *Vibrio cholerae (V. cholerae)*, and Pepper Mild Mottle Virus (PMMoV) as a fecal indicator.

**Findings:** Enteric viruses, non-O1/O139 *V. cholerae,* and PMMoV were consistently detected across all drainage channels and WWTP influents. Concentrations were generally lower in drainage than WWTP influents, except for non-O1/O139 *V. cholerae*. When normalized by PMMoV, concentrations across most drainage channels were comparable to WWTP influents, although comparability varied by target and location. Both concentrations and PMMoV-normalized concentrations varied across sampling locations. Trends between campaigns varied by pathogen target and were not explained by micro-catchment characteristic. Influenza A virus was the most frequently detected respiratory virus (8-23% in drainage channels; 3-10% in WWTP influents), while SARS-CoV-2, Influenza B, and RSV were rarely detected.

**Interpretation:** PMMoV-normalized concentrations across most micro-catchments were comparable to WWTP influents, with spatial heterogeneity implying neighborhood-level differences in disease prevalence. Catchment-delineated drainage surveillance has potential to offer spatially resolved public health information in non-sewered settings comparable to sewer-based wastewater monitoring.

**Funding:** Eawag Discretionary Funding

## Introduction

Robust surveillance systems are cornerstones of public health practice, providing timely and actionable data that support the early detection, monitoring, and control of infectious disease transmission.^1^ Primarily, infectious disease surveillance is based on aggregation of clinical data, which has notable challenges, including underreporting associated with asymptomatic or mildly symptomatic infections^2^, incentives that may discourage complete case reporting^3^, and limited access to health facilities in resource-constrained settings.^4^ These limitations are particularly acute in low- and middle-income countries (LMICs), where surveillance is often fragmented and insufficient to guide timely public health responses.^5^ Wastewater and environmental surveillance (WES) has effectively complemented clinical surveillance, enabling anonymous insights into the health of populations in a non-invasive, unbiased, and cost-effective approach.^6^ Pathogen dynamics in wastewater influents at centralized wastewater treatment plants (WWTPs) have been shown to reflect disease trends within the catchment’s population.^7^ Beyond trend monitoring, WES has triggered public health actions. For example, poliovirus detection in sewage and surface waters has directly triggered vaccination campaigns, demonstrating the actionable public health value of WES.^8^ Despite the promise of WES in disease control, a critical limitation is the absence of a defined relationship between the sampling location and the surveyed population in non-sewered settings. Unlike centralized WWTPs, in non-sewered areas, sampling sites are rarely linked to a known upstream community. Identifying the population that contributes to samples could ensure non-sewered WES is as actionable as sewer-based surveillance.

Nearly half of the global population relies on non-sewered sanitation.^9^ In urban areas of LMICs, it is often higher: for 60 to 90% of residents, wastewater is stored in pits and tanks (‘fecal sludge’) before road-based transport to treatment facilities.^10^ With rapid urbanization, the non-sewered population is growing faster than the population with access to sewers.^11^ Pits and tanks in urban areas are often informal and self-constructed, emptying is costly and logistically complex, and as a result, containments frequently overflow or are discharged directly into the urban environment. Approximately half of the fecal waste from non-sewered sanitation in urban areas has been estimated to leak into the environment, commonly entering surface waters.^12^

In urban areas where sewered and non-sewered sanitation coexist, drainage channels receive inputs from multiple pathogen sources (**Fig. 1**). The pathogen load from non-sewered wastewater will potentially depend on how diluted the wastewater is (e.g., flush type, level of water supply service, greywater management). It is also governed by how well contained it is (e.g., unlined, partially lined, or fully sealed pits or tanks).

**Fig. 1.**
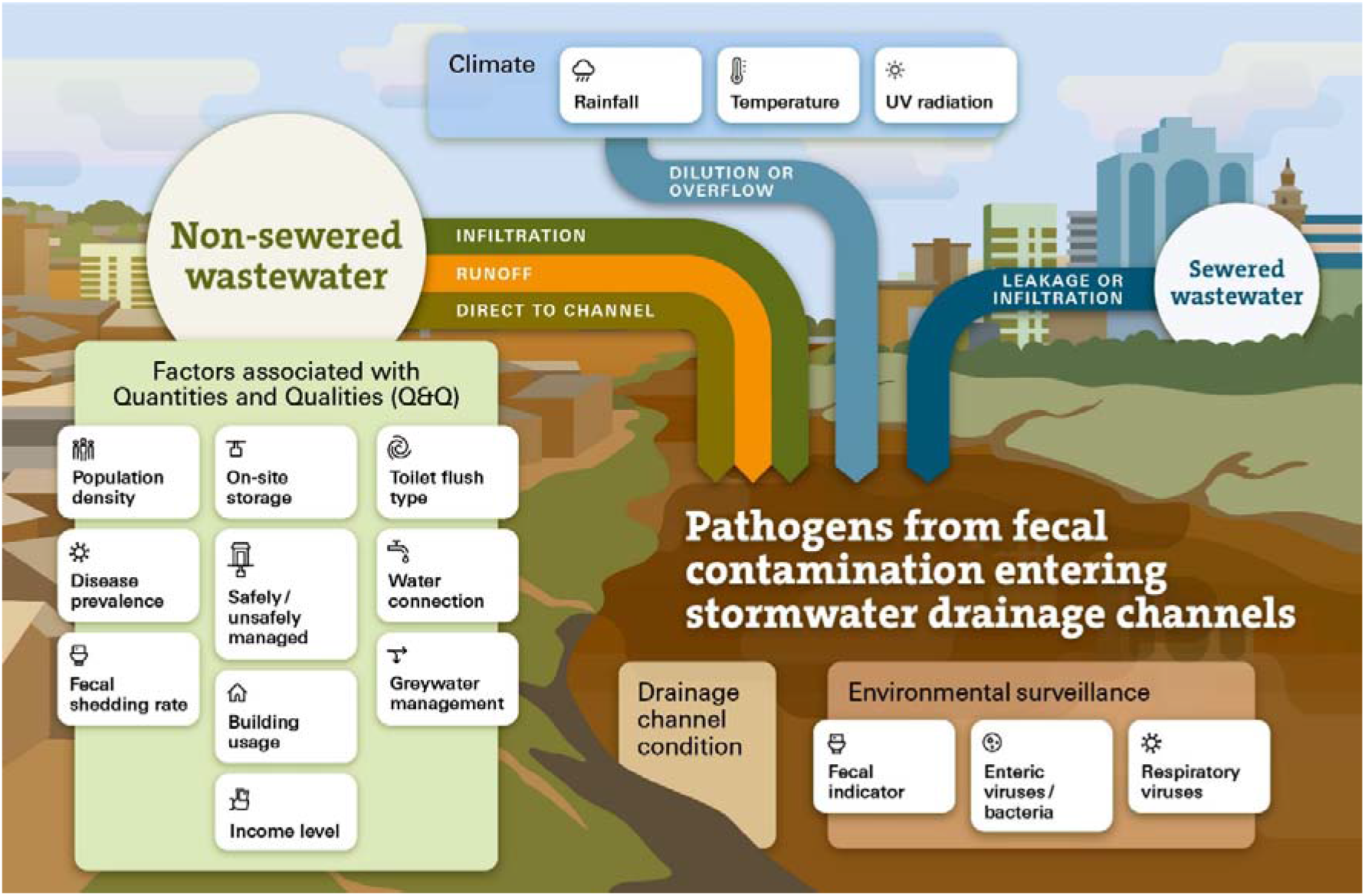
Conceptual model of pathogen transport via wastewater from non-sewered and sewered sanitation into urban drainage systems. Multiple components can directly or indirectly influence wastewater inputs to the drainage channel, collectively shaping pathogen levels detected therein. Non-sewered wastewater enters via infiltration, surface runoff, or direct discharge, with wastewater quantity and quality governed by on-site storage type, management practices, and household technical factors, while pathogen levels are further determined by population density, disease prevalence, and fecal shedding rates. Sewered wastewater contributes through leakage or overflow. Meteorological conditions impact concentrations through dilution or inactivation. Fecal indicators, enteric viruses and bacteria, and respiratory viruses serve as surveillance targets.

Pathogen levels in drainage channels reflect upstream generation, governed by population density, disease prevalence, and fecal shedding rates, as well as wastewater quantity and quality, for which building usage and income level can be statistical predictors of total solids concentrations.^14^ These levels are further impacted at the channel level by filtration through unlined channel walls depending on local soil characteristics^15^, and by meteorological conditions through rainfall-driven dilution or overflow, temperature impacting persistence, and solar UV inactivation.^16^

Pathogens and fecal indicators have previously been detected in drainage channels in cities with non-sewered sanitation.^17^ However, these detections have rarely been linked to spatially defined upstream communities. A key question is whether systematically delineating catchments above drainage channel sampling points can provide pathogen signals of comparable public health value to those from centralized WWTP influents.

In this study, we evaluated the potential of samples from urban drainage channels as a multi-pathogen WES matrix in Kampala, Uganda. We monitored enteric and respiratory pathogens, and Pepper Mild Mottle Virus (PMMoV) as a human fecal indicator due to its high prevalence and stability in human feces.^18^ We defined catchments and sampled from ten drainage channels, and in parallel sampled two WWTP influents as a reference. We addressed two main questions: first, whether spatially defined drainage channel catchments can provide community-level pathogen signals comparable to WWTP influents, assessed through pathogen detectability, concentration comparison, PMMoV normalization, and relationship to catchment characteristics; and second, what the observed spatiotemporal patterns imply for sample site selection of drainage-based WES systems. The findings aim to inform inclusive surveillance strategies for urban populations underserved by sewer-based surveillance.

## Methods

### Study site: Kampala, Uganda

The population of Kampala is estimated at 1·8 million.^19^ Based on the Excreta Flow Diagram (SFD)^20^, 15% are connected to a sewer network while 85% are served by non-sewered sanitation with road-based transport to fecal sludge treatment plants (FSTPs). Lubigi and Nakivubo are both WWTPs and FSTPs and Nalukolongo is a FSTP (**Fig. 2a**), An estimated 29% of total excreta in Kampala is unsafely managed, including 13% of sewered wastewater and 38% of non-sewered wastewater.^20^

**Fig. 2.**
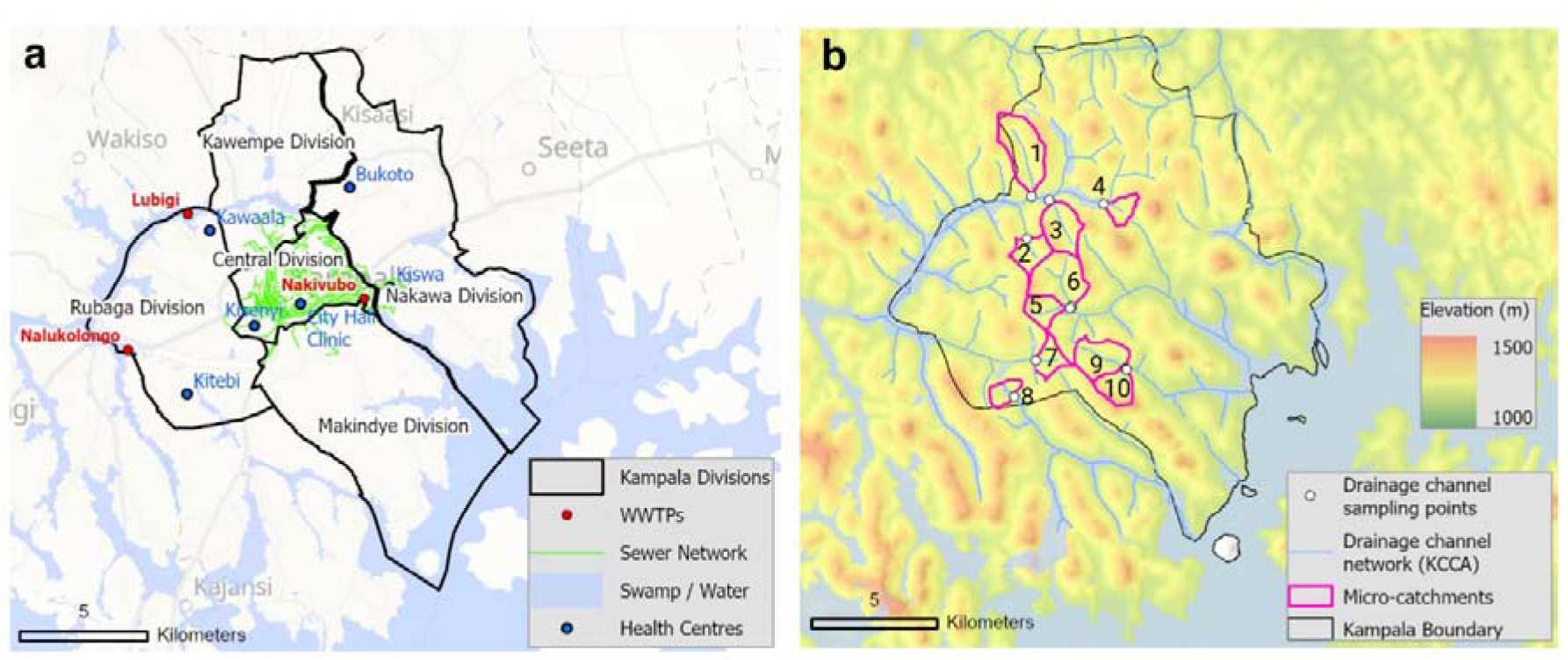
Geospatial overview of the study area in Kampala, Uganda. (a) Five administrative divisions of Kampala with the centralized sewer network, the locations of three wastewater treatment plants (WWTPs), Lubigi, Nakivubo, and Nalukolongo (sampling was conducted at Lubigi and Nakivubo in this study), and the locations of health centers, Kawaala, Bukoto, Kisenyi, City Hall Clinic, Kiswa, and Kitebi. (b) Delineated micro-catchments of the ten drainage channel sampling points: #1-10, derived from a digital elevation model (DEM) overlaid on elevation data. Note that the sampling points #5 and #6 were shifted downstream during the second sampling campaign (#5’ and #6’) (**Fig. S1**). Background map sources: Esri, TomTom, Gamin, FAO, NOAA, USGS, © OpenStreetMap Contributors, GIS User Community, CGIAR, USGS.

### Sampling site selection and micro-catchment delineation

The map of the drainage system for the city was obtained from Kampala Capital City Authority (KCCA) and used to identify candidate sampling locations at the lowest elevation point of each delineated micro- catchment, ensuring that flow at the sampling point reflected the area upstream. Each micro-catchment encompassed between 10,000 and 50,000 people, a scale chosen to enable community-level surveillance while limiting demographic heterogeneity. Twenty-two candidate micro-catchments were delineated across the Kampala drainage network using the *Watershed Delineation* tool in Arc Hydro (ArcGIS Pro, ESRI, v.3.6.2), with a 5-m resolution digital elevation model (DEM) as input. For each micro-catchment, population was estimated using 100-m gridded data.^21^

An expert group of practitioners and researchers from KCCA, Makerere University, and Eawag evaluated the 22 candidate micro-catchments across multiple criteria, including sanitation conditions, drainage channel condition, socioeconomic characteristics, population coverage, with expected pathogen levels determined through expert knowledge (**Annex 1**). Field validation was subsequently conducted at all 22 micro-catchments to verify the presence of sufficient flow, document drainage channel condition and geospatial location at division and parish level. Ten micro-catchments, #1-10, were selected to ensure representative distribution of catchment area (0·95 to 3·83 km^2^), population (12,885 to 44,146), and drainage channel condition (lined or unlined) (**Fig. 2b** and **Table 1**). Sampling points #5 and #6 were shifted approximately 0·6 km downstream during the second sampling campaign due to construction limiting access to the drainage channel and are annotated as #5’ and #6’ (**Fig. S1**).

**Table 1.** Characteristics of ten micro-catchments included in this study. Catchment area and population of delineated micro-catchments, #1-10. Drainage channel condition refers to the physical lining status of the drainage channel, where lined channels have walls and are constructed of concrete or masonry, and unlined channels are earthen, allowing direct contact between channel water and surrounding soil. Administrative division and parish were recorded from field observations and geospatial data. Sampling points #5 and #6 were located in central market areas. During the second sampling campaign, both points were shifted downstream due to land-use change and difficulty accessing the drainage channel (**Fig S2**). #5’ became the combined micro- catchments of #5 and #6 and #6’ is a new micro-catchment.

|  | Micro-catchment area<br>(km<sup>2</sup>) | Population | Drainage channel<br>condition | Division | Parish |
| --- | --- | --- | --- | --- | --- |
| #1 | 3.83 | 44,146 | Lined | Kawempe | Bwaise |
| #2 | 1.57 | 17,942 | Unlined | Kawempe | Makerere<br>Kikoni/Kawaala |
| #3 | 2.54 | 30,117 | Lined | Kawempe | Makerere Kavule/<br>Mulago |
| #4 | 1.47 | 18,576 | Unlined | Nakawa/Central | Kamwokya |
| #5 | 1.41 | 17,356 | Lined | Central | Kisenyi |
| #5' | 7.25 | 84,494 | Lined | Central | Kisenyi |
| #6 | 3.66 | 41,906 | Lined | Central | Kisenyi |
| #6' | 0.71 | 9,521 | Lined | Makindye | Kabalagala |
| #7 | 1.29 | 14,718 | Unlined | Rubaga | Nalukolongo/<br>Ndeeba |
| #8 | 0.95 | 12,885 | Lined | Rubaga | Wankulukuku |
| #9 | 2.66 | 34,792 | Lined | Makindye | Nsambya |
| #10 | 1.65 | 18,453 | Unlined | Makindye | Nsambya |

### Environmental sampling

Samples were conducted during two campaigns: the first between 17 and 28 March 2025 (ten sampling days) and the second between 15 September and 10 October 2025 (16 sampling days), covering ten drainage channel sampling points and two WWTP influents. The first campaign yielded 118 samples (98 drainage channels, 20 WWTP influents) and the second 190 samples (156 drainage channels, 34 WWTP influents). At drainage channel sampling points, 200 mL grab samples were collected between 9:00 AM and 2:00 PM in single-use disposable HDPE containers, immediately stored on ice, and transported to the laboratory at Makerere University within five hours. Concurrently, the volumetric flow rate (m^3^/day) was recorded at each sampling event (**Text S1**). At Lubigi and Nakivubo WWTPs, influent samples were grab-sampled each morning between 9:00 and 11:00 AM and stored onsite at 4°C until weekly retrieval and transport on ice to the laboratory.

### Sample concentration and DNA/RNA extraction

Each grab sample was aliquoted to 50 mL, treated with MgCl_2_ (25 mM final concentration), and vacuum-filtered through a 0.45-µm mixed cellulose ester membrane filter (**Text S2**).^22^ DNA and RNA were co-extracted from filters using AllPrep PowerFecal Pro DNA/RNA Kit (Qiagen, Cat. No. 80254) and eluted separately in 100 µL of DNase/RNase-free water. Extracts were purified, 3× diluted, and stored at −80 °C until being transported on dry ice to Eawag, Switzerland for analysis. Extraction blanks using DNase/RNase-free water were processed in parallel with each extraction day.

### Digital PCR assays for pathogen quantification

Multiple pathogens were quantified in gene copies in sample volume (gc/mL) using a one-step digital RT-PCR or PCR assays on the naica® PCR platform (Stilla Technologies, Villejuif, France) following the Minimum Information for Publication of Quantitative Digital PCR Experiment (dMIQE) guidelines^23^ (**Annex 2**). Enteric targets included Norovirus GI, Norovirus GII, Rotavirus, and *Vibrio cholerae* (*V. cholerae*), the latter targeting both the species-specific marker and serogroup markers for O1 and O139. Respiratory viruses included SARS-CoV-2, Influenza A and B viruses, Respiratory syncytial virus (RSV). PMMoV was also quantified. All reactions were performed in duplicate with positive and no-template controls using previously published primer and probe sequences with some modification for some targets (**Table S1**) and thermocycling conditions (**Text S3**).^24–31^ A sample was considered detectable if three or more positive droplets were observed per well, corresponding to ∼8.1 gc/mL. The limit of blank (LOB) was defined as five positive droplets across all assays; one extract blank exceeded the LOB for Rotavirus on 22 September 2025 and Rotavirus data extracted on that day was excluded from analysis (**Table S2**). A subset of samples tested for inhibition, showed limited extent (**Table S3**).

### Culture and Physicochemical analyses

*E. coli* was enumerated using CompactDry® EC plate (Shimadzu) and total suspended solids (TSS) were calculated by a filtration method (**Text S4**).

### Meteorological data

Daily rainfall (mm) during the sampling periods were obtained from Uganda National Meteorological Authority (data available on request). Two sampling campaigns were designed to capture contrasting rainfall conditions: March campaign coincided with the first rainy season (average 12.3 mm/day over the 12-day sampling period), and September-October campaign with the second rainy season of lower intensity (average 4.0 mm/day over 26-day sampling period) (**Table S4**).

### Clinical data

Clinical surveillance data at six health centers, Kawala, Kitebi, Kiswa, Bukoto, Kisenyi, and City Hall located near sampling points were collected from the Ministry of Health to help contextualize WES results. Data included the weekly number of cases of diarrheal and respiratory diseases reported between January 2023 and December 2025.

### Ethics approval

The protocols for collecting and processing samples were approved by the Uganda National Council for Science and Technology (UNCST) (approval no. HS5809ES), the Vector Control Division Research Ethics Committee in Uganda (approval no. VCDR-2024-65) and the Eawag Ethical Review Committee.

## Results

### Multi-pathogen and Fecal Indicator Monitoring Across Drainage Channels

During the sampling periods, enteric viruses (Norovirus GI and GII, Rotavirus), non-O1/O139 *V. cholerae*, and PMMoV were consistently detected in both drainage channels and WWTP influents (**Fig. 3a**). Pooled across both sampling campaigns, PMMoV and Rotavirus showed the highest median concentrations in both drainage channels (3·7 and 3·8 log_10_ gc/mL) and WWTP influents (4·3 and 4·6 log_10_ gc/mL) (**Table 2**).

**Fig. 3.**
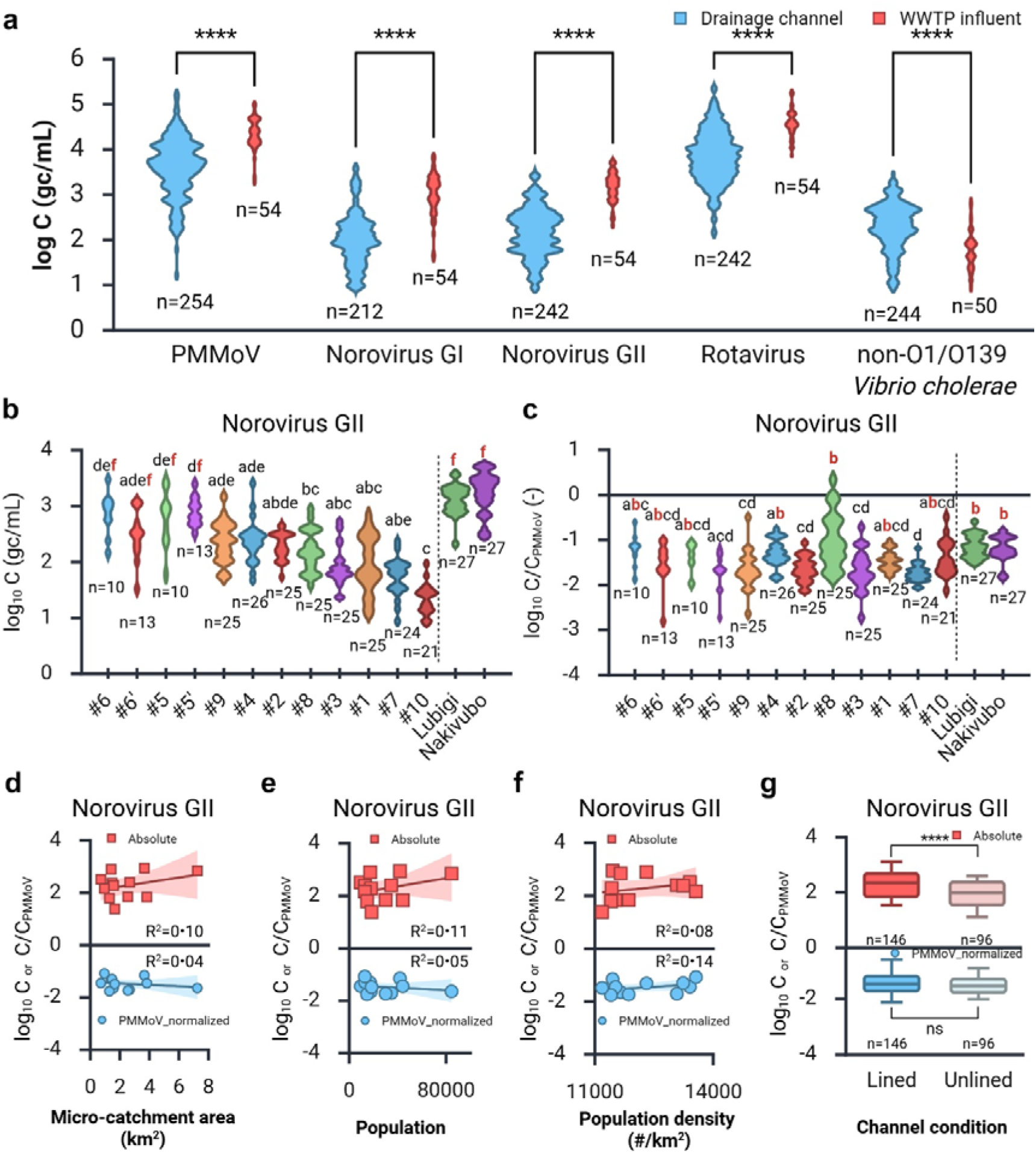
Multi-target monitoring of fecal indicator and enteric pathogens in drainage channel and wastewater influent to the centralized treatment plants (WWTPs) (a) Violin plots of log_10_ concentrations (gc/mL) of PMMoV, enteric viruses (Norovirus GI and GII, and Rotavirus) and non-O1/O139 *Vibrio cholerae* (*V. cholerae*) across all drainage channel and WWTP influent sampling points. Spatial variation of log_10_ concentrations (b) and PMMoV-normalized concentrations (c) of Norovirus GII across ten individual drainage channel sampling points, #1-10, and two WWTP influents, Lubigi and Nakivubo, ordered from left to right by ascending median concentration for drainage channels, with WWTP influents on the right. Violin plots are scaled by count with light smoothing. (d-g) Association between concentrations and micro-catchment characteristics. (d-f) Scatter plots of median log_10_ concentrations (upper panels) and PMMoV-normalized concentrations (lower panels) of Norovirus GII across micro-catchments by catchment area (km^2^) (d), population (e), and population density (#/km^2^) (f). The line and shaded region indicate best-fit linear regression with 95% confidence interval and R-square value. (g) Boxplot of log_10_ concentrations (upper) and PMMoV-normalized concentrations (lower) of Norovirus GII by drainage channel condition (lined vs. unlined), showing median and 5^th^-95^th^ percentile range. Statistical comparisons are indicated as follows: Two-way ANOVA with Bonferroni correction for (a) (**** p<0·0001; ns: not significant); Kruskal-Wallis test with Dunn’s pairwise multiple comparisons for (b, c). Group differences are summarized using compact letter display (CLD); letters shared between WWTP influents and drainage channels are highlighted in red; Mann-Whitney U test for (g).

**Table 2.** Median log_10_ concentrations (gc/mL) [25^th^, 75^th^ percentile] and PMMoV-normalized concentration (log_10_ gc/mL target per log_10_ gc/mL PMMoV) of enteric viruses and non-O1/O139 *V. cholerae* across ten drainage channel and two wastewater treatment plant (WWTP) influent sampling points during two campaigns. n indicates the number of samples. Between-campaign differences were compared using two-way ANOVA with Bonferroni correction (**** p<0·0001; *** p<0·001; ** p<0·01 ns: not significant).

| Sampling period | Drainage channel |  |  |  |  |  |  | WWTP influent |  |  |  |  |  |  |
| --- | --- | --- | --- | --- | --- | --- | --- | --- | --- | --- | --- | --- | --- | --- |
|  | Total | n | March | n | Sep-Oct | n | Campaign difference | Total | n | March | n | Sep-Oct | n | Campaign difference |
| PMMoV | 3.7 [3.2, 4.1] | 254 | 3.4 [2.9, 4.0] | 98 | 3.8 [3.4, 4.1] | 156 | **** | 4.3 [4.2, 4.6] | 54 | 4.2 [4.1, 4.5] | 20 | 4.4 [4.2, 4.7] | 34 | ns |
| Norovirus GI | 2.0 [1.6, 2.3] | 212 | 1.6 [1.2, 1.9] | 66 | 2.1 [1.9, 2.4] | 146 | **** | 3.1 [2.8, 3.4] | 54 | 2.8 [2.4, 2.9] | 20 | 3.2 [3.1, 3.4] | 34 | *** |
| Norovirus GII | 2.2 [1.8, 2.6] | 242 | 2.2 [1.7, 2.6] | 90 | 2.2 [1.8, 2.5] | 152 | ns | 3.2 [2.9, 3.4] | 54 | 3.3 [3.2, 3.6] | 20 | 3.1 [2.9, 3.3] | 34 | ns |
| Rotavirus | 3.8 [3.5, 4.2] | 242 | 3.6 [3.2, 4.0] | 98 | 4.0 [3.6, 4.4] | 144 | **** | 4.6 [4.4, 4.8] | 54 | 4.6 [4.5, 4.8] | 20 | 4.6 [4.4, 4.8] | 34 | ns |
| non-O1/O139 *V. cholerae* | 2.4 [2.0, 2.7] | 244 | 2.2 [1.8, 2.6] | 94 | 2.4 [2.1, 2.7] | 150 | ** | 1.7 [1.5, 2.0] | 50 | 1.8 [1.3, 2.1] | 18 | 1.7 [1.5, 1.9] | 32 | ns |
| PMMoV-normalized |  |  |  |  |  |  |  |  |  |  |  |  |  |  |
| Norovirus GI | -1.8 [-2.0, -1.5] | 212 | -2.0 [-2.3, -1.7] | 66 | -1.6 [-1.9, -1.4] | 146 | **** | -1.3 [-1.6, -1.1] | 54 | -1.5 [-1.7, -1.4] | 20 | -1.2 [-1.4, -1.0] | 34 | ** |
| Norovirus GII | -1.5 [-1.8, -1.2] | 242 | -1.3 [-1.6, -1.0] | 90 | -1.6 [-1.8, -1.4] | 152 | **** | -1.2 [-1.4, -0.9] | 54 | -0.9 [-1.0, -0.8] | 20 | -1.3 [-1.4, -1.2] | 34 | *** |
| Rotavirus | 0.2 [-0.1, 0.6] | 242 | 0.2 [-0.2, 0.6] | 98 | 0.2 [-0.1, 0.6] | 144 | ns | 0.1 [-0.1, 0.5] | 54 | 0.3 [0.0, 0.5] | 20 | 0.0 [-0.2, 0.5] | 34 | ns |
| non-O1/O139 *V. cholerae* | -1.4 [-1.7, -1.0] | 242 | -1.3 [-1.6, -0.8] | 92 | -1.5 [-1.7, -1.2] | 150 | ** | -2.6 [-3.0, -2.4] | 50 | -2.5 [-2.6, -2.3] | 18 | -2.7 [-3.0, -2.4] | 32 | ns |

Overall, concentrations were significantly higher in WWTP influents than in drainage channels across all targets, as well as other fecal indicators, *E. coli* (CFU/mL) and TSS (mg/L) (**Fig. S2**), except for non- O1/O139 *V. cholerae*, which were significantly higher in drainage channels than WWTP influents (**Fig. 3a**). For *V. cholerae*, in addition to the species-specific *ompW* marker, the *O139rfb* serogroup marker was detected in three drainage channel samples, indicating sporadic presence of the toxigenic O139 serogroup alongside the predominantly non-O1/O139 signal.

Spatial variation in concentrations across drainage channels and WWTP influents was further examined (Norovirus GII in **Fig. 3b**; Norovirus GI, Rotavirus, and non-O1/O139 *V. cholerae* in **Fig. S3**). For Norovirus GII, distinct spatial differences were observed across drainage channels, with both WWTP influents showing the highest concentrations. Two drainage channel sampling points located in the central market area, #5 and #6, and #5’ and #6’, reached comparable levels to WWTP influents.

Following PMMoV normalization, a larger subset of drainage channel sampling points showed concentrations comparable to WWTP influents (Norovirus GII in **Fig. 3c**; remaining targets in **Fig. S3**). For Norovirus GII, concentrations at seven sampling points were comparable to the WWTP influents while remaining five points were significantly lower. Daily pathogen load normalized by micro-catchment population and flow rate (gc/person/day) was also examined as a complementary normalization approach (**Fig. S4**). The spatial pattern was broadly consistent with PMMoV-normalization, with most drainage channels comparable across micro-catchments.

Micro-catchment characteristics including area, population, population density, and drainage channel conditions were examined for their association with pathogen concentrations (Norovirus GII in **Fig. 3d-f**; remaining targets in **Fig. S5**) and were not significantly correlated with either concentrations or PMMoV- normalized concentrations. Lined drainage channels showed higher concentrations than unlined channels, however, no significant difference was observed in PMMoV-normalized concentration between the two channel conditions.

Concentrations were also compared between two sampling campaigns (**Table 2**). PMMoV-normalized concentrations differed significantly between campaigns for Norovirus GI and GII in both drainage channels and WWTP influent, with Norovirus GI higher in September/October and Norovirus GII higher in March. Rotavirus was not significantly different between campaigns in either drainage channels or WWTP, and non-O1/O139 *V. cholerae* was significantly different between campaigns in drainage channels but not WWTPs. Absolute concentrations differed significantly between campaigns for PMMoV, Norovirus GI, Rotavirus, and non- O1/O139 *V. cholerae* in drainage channels and for Norovirus GI only in WWTP influents, with all observing higher concentrations in September/October. Norovirus GII showed no significant difference between-campaign difference in either matrix.

### Temporal Dynamics of Pathogens in Drainage channels and WWTP influents

Temporal dynamics of PMMoV-normalized concentrations were observed across drainage channels and WWTP influents (Rotavirus in **Fig. 4a**; Norovirus GI and GII, and non-O1/O139 *V. cholerae* in **Fig. S6**).

**Fig. 4.**
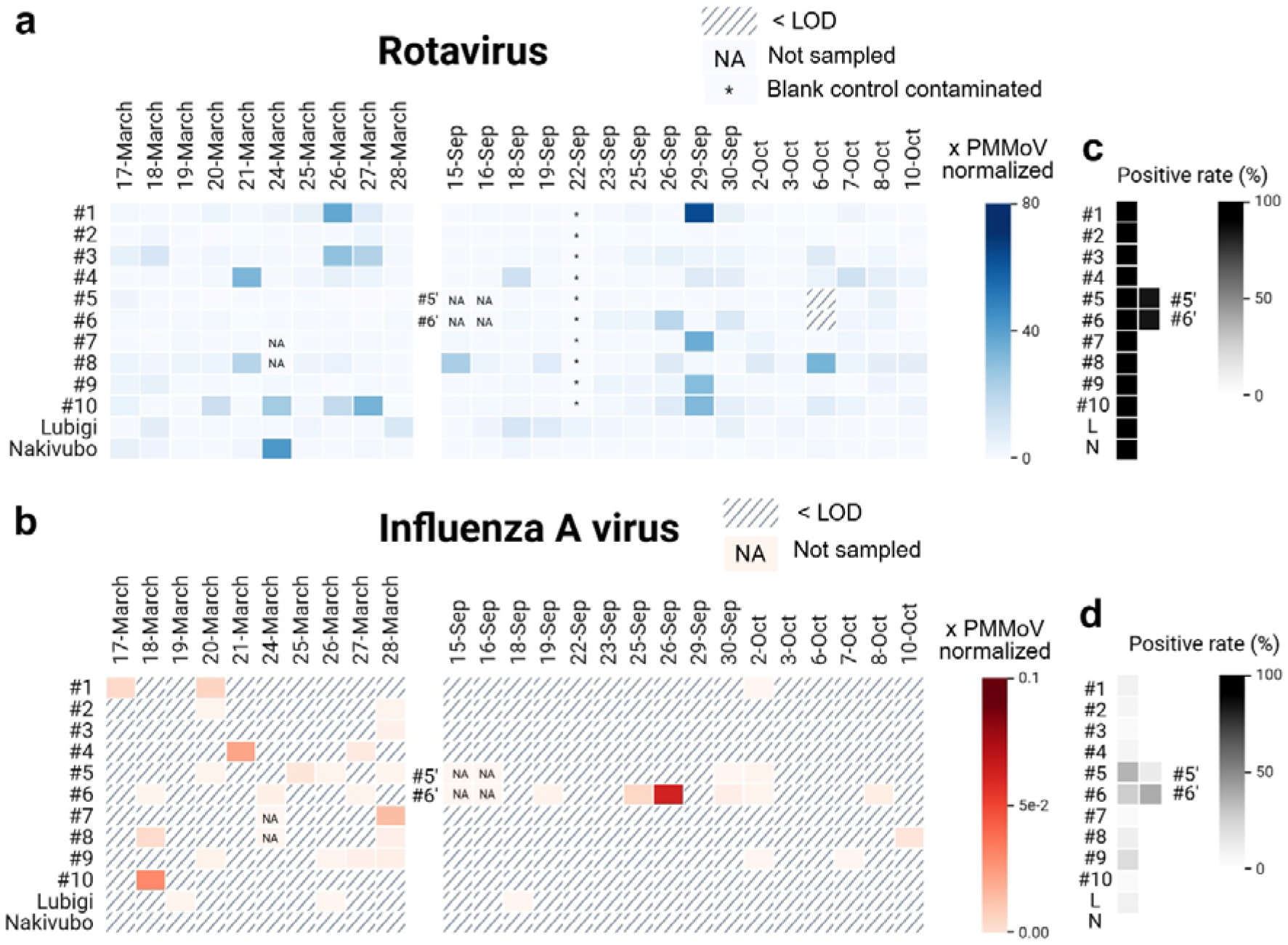
Temporal dynamics of PMMoV-normalized pathogen concentrations in drainage channels and wastewater influents to the centralized treatment plants (WWTPs) Heatmaps of PMMoV-normalized concentrations of Rotavirus (a) and Influenza A virus (b) across ten drainage channel sampling points, #1-10, and two WWTP influents, Lubigi and Nakivubo, over two sampling campaigns from 17 to 28 March 2025 and from 15 September to 10 October 2025. Hatched cells indicate concentrations below the limit of detection (LOD); “NA” indicates missing samples. Asterisks on specific date indicate blank control contamination, and the corresponding samples were excluded from analysis. Heat maps of positive rate of samples (%) for Rotavirus (c) and Influenza A virus (d) are shown for individual drainage channel sampling point and each WWTP influent, annotated L (Lubigi) and N (Nakivubo).

Positive rates were lowest at #10 for Norovirus GI (38%), Norovirus GII (81%), non-O1/O139 *V. cholerae* (84%). Rotavirus was always detected throughout both campaigns, except for 6 October at #5’ and #6’ following a flooding event (**Fig. S7**).

Among respiratory targets, Influenza A virus was sporadically (**Fig. 4b**), with higher positive rates in March than in September-October in both drainage channels (23% vs. 8%) and WWTP influents (10% vs. 3%). Notably higher rates were observed at #5 (40%) and #6’ (43%). SARS-CoV-2 was detected in drainage channels (4%) and WWTP influents (25%) during March campaign, but only in WWTP influents (6%) during September-October campaign. Influenza B virus and RSV were rarely detected, with single positives in WWTP influents and drainage channels respectively during March campaign, and no detections in either matrix during September-October campaign.

### Disease incidence trends based on clinical surveillance

The detection of respiratory and diarrheal diseases through clinical surveillance at health centers varied over time and between facilities and suggests almost continuous transmission of both influenza-like illness and dysentery between 2023 and 2025 (**Fig. S8**). A total of 4,819 cases of influenza-like illness were reported across three health centers located near sampling points for three years, while other respiratory diseases were less frequent, with 48 cases of severe pneumonia among children under five years reported by two health centers, and 14 cases of SARS reported by two health centers (**Table S5**). Dysentery was reported by five health centers, for a total of 193 cases over three years, and 109 cases of diarrhea with dehydration were recorded among children under five years across two health centers. No marked differences in diarrheal or respiratory illnesses were observed between the two campaign periods. No cholera and no Covid-19 case were reported during the study period. The City Hall Clinic did not report any case of the diseases listed above.

## Discussion

This study demonstrates that catchment-delineated urban drainage channels can provide spatially resolved, community-level pathogen signals, and that pathogen detection rates and normalized concentrations are comparable to sewer-based surveillance. Enteric pathogens and fecal indicators were reliably detectable across all drainage channel sampling points, consistent with the findings from drainage channels and surface waters in other cities, including Kampala.^32,33^ Concentrations were significantly higher in WWTP influents than in drainage channels, reflecting the concentrated wastewater transported in the sewer compared to the diluted and heterogeneous inputs into the drainage channels. Notably, Influenza A virus was detected more frequently in drainage channels than in WWTP influents, suggesting that drainage channels may capture respiratory virus circulation at the community level that is not fully reflected in sewer-based surveillance. The exception was non- O1/O139 *V. cholerae*, which was significantly higher in drainage channels, likely reflecting its known environmental reservoir in urban surface waters rather than exclusive fecal origin, as previously reported in Ugandan surface water.^34^

Comparability of WES in non-sewered to sewered settings highlights the potential opportunities for WES to support pathogen surveillance, particularly in regions with limited disease surveillance or where use of clinical data in surveillance may be insufficient due to challenges identified earlier. Clinical surveillance data from health facilities in Kampala confirmed a persistent burden of respiratory and diarrheal disease in the study areas. However, several factors limit the feasibility of direct comparison between drainage channel pathogen signals and clinical surveillance data. These include the short duration of sampling campaigns, differences in spatial scales (micro-catchments vs. health centers), potentially diverse care-seeking behaviors, and the reliance of clinical surveillance on syndromic rather than pathogen-specific diagnostics. Further studies should assess the opportunity for WES to provide consistent and spatially resolved community-level disease information that is representative of community health and complementary to clinical surveillance.

Pathogen concentrations varied substantially across drainage sampling points, but these differences were not significantly associated with micro-catchment area, population, or population density across all targets. While lined drainage channels showed higher concentrations than unlined channels, possibly reflecting partitioning of pathogens onto sediment and soil in unlined channels.^35^ PMMoV-normalized concentrations did not significantly differ between the two drainage channel conditions. Notably, PMMoV-normalization to account for fecal load revealed that most drainage channels had comparable pathogen signals to WWTP influents, suggesting that drainage channels reflect community-level disease circulation in an analogous fashion to sewer-based surveillance. A subset of the drainage channels showed significantly lower normalized signals, potentially indicating lower pathogen prevalence in the corresponding upstream communities. Daily pathogen load normalized by micro-catchment population and flow rates showed a broadly similar spatial pattern to PMMoV normalization. As illustrated in the conceptual framework (**Fig. 1**), the absence of strong associations with individual micro-catchment characteristics underscores that pathogen levels in drainage channels are governed by a combination of interconnected factors across the sanitation service chain, highlighting the need for integrated sanitation data at the neighborhood scale to interpret spatial variation.

Pathogen concentrations in drainage channels had mixed patterns between the campaigns. The conceptual framework (**Fig. 1**) highlights the competing meteorological effects, higher rainfall likely increased volumetric flow diluting pathogen concentrations, while more episodic rainfall may have mobilized wastewater through increased infiltration and overflow of on-site containments, driving concentrations upwards. Seasonal patterns in Kampala drainage channels and similar urban settings in Uganda appear inconsistent in the literature. Sadik et al.^32^ reported higher fecal contamination in Kampala drainage channels during the wet season, while Ronoh et al.^36^ found higher *E. coli* in drainage channels during the dry season in a low-income Ugandan settlement, suggesting local sanitation conditions and rainfall patterns interact in ways that preclude simple seasonal generalizations. In an extreme case, a flooding event at channel #5’ on October 6 caused a marked drop in all target concentrations, with full recovery within one day (**Fig. S7**), highlighting that extreme rainfall and flooding events can substantially affect drainage channel pathogen signals. Since both campaigns fell within wet seasons of differing intensity rather than representing a true wet-dry contrast, interpretation of between- campaign differences is limited, and future sampling spanning distinct dry and wet seasons is needed to fully characterize meteorological drivers of pathogen dynamics in urban drainage channels.

Among respiratory viruses, Influenza A virus was sporadically but recurrently detected across both drainage channels and WWTP influents, with more frequent detection and higher concentrations observed in drainage channels. These detections illustrate the potential utility of drainage-based surveillance for monitoring respiratory pathogens, with opportunities for genomic characterization of circulating strains, as demonstrated for Influenza A virus in wastewater.^37^ The higher positive rates observed at specific drainage channel points suggest that certain micro-catchments may experience higher respiratory virus circulation, further supporting the value of spatially resolved drainage-based surveillance for identifying communities with disproportionate infectious disease circulation. Unlike sewer-based surveillance, where population equivalents from a sewer-shed can be estimated from flow and fecal strength, drainage channel catchments require explicit spatial delineation and population mapping to link sampling locations to defined communities. However, static population mapping does not capture population mobility. The higher pathogen concentrations observed in central market areas (#5 and #6) may reflect disproportionate fecal inputs from commuting daytime populations not captured by residential counts, highlighting that population equivalents accounting for dynamic population fluxes may be needed to accurately characterize the fecal input to channels.

Drainage channels in Kampala carry continuous flow year-round, suggesting drainage-based WES could support routine longitudinal surveillance. These findings support the development of multi-panel WES in urban drainage channels as a complement, consistent with the World Health Organization call for expanding WES beyond centralized treatment infrastructure.^38^ A key methodological advance demonstrated here is the delineation of micro-catchments as population-defined surveillance units analogous to sewer-based surveillance, enabling attribution of environmental pathogen signals to known upstream communities. Spatially resolved sampling strategies, informed by catchment population equivalents, income level, and sanitation coverage, could further maximize surveillance value and enable targeted monitoring of neighborhoods with high disease circulation. Realizing this potential will require investment in laboratory capacity, stakeholder engagement, and the democratization of WES infrastructure to ensure that communities currently invisible to conventional surveillance systems are included in global health monitoring efforts.^39^

Beyond the surveillance utility of urban drainage, the consistent detection of high pathogen levels underscores the public health burden associated with unsafe management of wastewater in urban areas. Drainage channels in cities with non-sewered sanitation create opportunities for human exposure to fecal pathogens. This dichotomy, that drainage channels are both a surveillance matrix and a direct exposure pathway, offers a more targeted basis for prioritizing sanitation infrastructure investments to reduce pathogen exposure while leveraging drainage channels for community health monitoring.

## Supporting information

Annex 1. Group discussion and field validation

Annex 2. dMIQE

## Associated Content

### Supplementary Information

**Text S1** Measurement of volumetric flow rate of drainage channels.

**Text S2** Sample concentration and DNA/RNA extraction.

**Text S3** Digital PCR assays for pathogen quantification.

**Text S4** *E. coli* enumeration and physicochemical analysis

**Fig. S1** Micro-catchments from two drainage channel sampling points, #5 and 6, during two sampling campaigns in March (a) and September/October (b).

**Fig. S2** Violin plots of log_10_ concentrations of *E. coli* and total suspended solids (TSS) across ten drainage channel sampling points and two WWTP influents.

**Fig. S3** Violin plots of log_10_ concentrations of Norovirus GI, Rotavirus, and non-O1/O139 *V. cholerae* and their PMMoV-normalized concentrations across ten drainage channel sampling points and two WWTP influents.

**Fig. S4** Violin plots of log_10_ daily pathogen load normalized by micro-catchment population and flow rate (gc/population/day) of Norovirus GI and GII, Rotavirus, and non-O1/O139 *V. cholerae* across ten drainage channel sampling points, #1-10.

**Fig. S5** Association between concentrations and micro-catchment characteristics.

**Fig. S6** Temporal dynamics of PMMoV-normalized pathogen concentrations in drainage channels and WWTP influents.

**Fig. S7** Impact of a flooding event on pathogen concentrations at drainage channel sampling point, #5’.

**Fig. S8** Monthly number of respiratory and diarrheal disease cases recorded through routine surveillance at official health centers located near sampling points.

**Table S1** Sequence of PCR primers and probes used in this study.

**Table S2** The number of positive droplets by dPCR assays on blank extracts.

**Table S3** Inhibition test against SARS-CoV-2 N1 on the subset of drainage channel and WWTP influent samples

**Table S4** Meteorological conditions during the two sampling campaigns in Kampala, Uganda.

**Table S5** Annual number of respiratory and diarrheal disease cases recorded through routine surveillance at official health centers located near sampling points.

## Acknowledgements

We declare no competing financial interest. The project is funded by Eawag Discretionary Funding. SK also received funding from the Eawag Discretionary Postdoctoral Fellowship. We acknowledge Rosi Siber, Dr. Jolinda de Korne-Elenbaas from Eawag, Mathias Kasirye, Emmanuel Wantono, Collin Irumba, and Davis Majara from Makerere University, National Water and Sewerage Corporation (NWSC), and Kampala Capital City Authority (KCCA). We would like to thank the Director General of Health Services and team at the Ministry of Health in Uganda for sharing clinical surveillance data for use in this study. Fig. 2, 3, and 4 were created in BioRender. Kang, S. (2025) https://BioRender.com/y1na0al

## CRediT Contributors

SK, TRJ, and LS were involved in the conceptualization of this study. SK, JZB, GM, FZM, AK, CBN, LS designed sampling plans and conducted field validation. SK, AK, GJSP performed experimental sample collection. CBN, KG provided clinical data and linked it to experimental data. SK and LS were responsible for project administration. SK, TRJ, and LS verified and visualized the data. TRJ and LS provided supervision and were responsible for funding acquisition. SK wrote the original draft of the manuscript. All authors critically reviewed the manuscript and were responsible for the final decision to submit this work for publication.

## Data Availability

All data have been deposited in the Eawag Research Data Institutional Collection (ERIC) at https://doi.org/10.25678/000GZS.

## Use of Artificial Intelligence

In the scope of this work, the AI language model Claude (Sonnet 4.6, accessible under claude.ai) was used for technical help in R and proofreading the manuscript. All outputs were verified by the authors.

## Notes

### Competing Interest Statement

The authors have declared no competing interest.

