## Supporting Information for "Wastewater and Environmental Surveillance in Cities with Sewered and Non-sewered Sanitation: Evidence from Kampala, Uganda"

**Electronic Supplementary Information**

Contents

### Text S1 Measurement of volumetric flow rate of drainage channels.

### Text S2 Sample concentration and DNA/RNA extraction.

### Text S3 Digital PCR assays for pathogen quantification.

### Text S4 *E. coli* enumeration and physicochemical analysis

**Fig. S1** Micro-catchments from two drainage channel sampling points, #5 and 6, during two sampling campaigns in March (a) and September/October (b).

**Fig. S2** Violin plots of log_10_ concentrations of *E. coli* and Total suspended solids (TSS) across ten drainage channel sampling points and two WWTP influents.

**Fig. S3** Violin plots of log_10_ concentrations of Norovirus GI, Rotavirus, and non-O1/O139 *V. cholerae* and their PMMoV-normalized concentrations across ten drainage channel sampling points and two WWTP influents.

**Fig. S4** Violin plots of log_10_ daily pathogen load normalized by micro-catchment population and flow rate (gc/population/day) of Norovirus GI and GII, Rotavirus, and non-O1/O139 *V. cholerae* across ten drainage channel sampling points, #1-10.

**Fig. S5** Association between concentrations and micro-catchment characteristics.

**Fig. S6** Temporal dynamics of PMMoV-normalized pathogen concentrations in drainage channels and WWTP influents.

**Fig. S7** Impact of a flooding event on pathogen concentrations at drainage channel sampling point, #5’.

**Fig. S8** Monthly number of respiratory and diarrheal disease cases recorded through routine surveillance at official health centers located near sampling points.

**Table S1** Sequence of PCR primers and probes used in this study.

**Table S2** The number of positive droplets by dPCR assays on blank extracts.

**Table S3** Inhibition test against SARS-CoV-2 N1 on the subset of drainage channel and WWTP influent samples

**Table S4** Meteorological conditions during the two sampling campaigns in Kampala, Uganda.

**Table S5** Annual number of respiratory and diarrheal disease cases recorded through routine surveillance at official health centers located near sampling points.

**Annex 1.** Group discussion and field validation.

**Annex 2**. dMIQE guidelines.

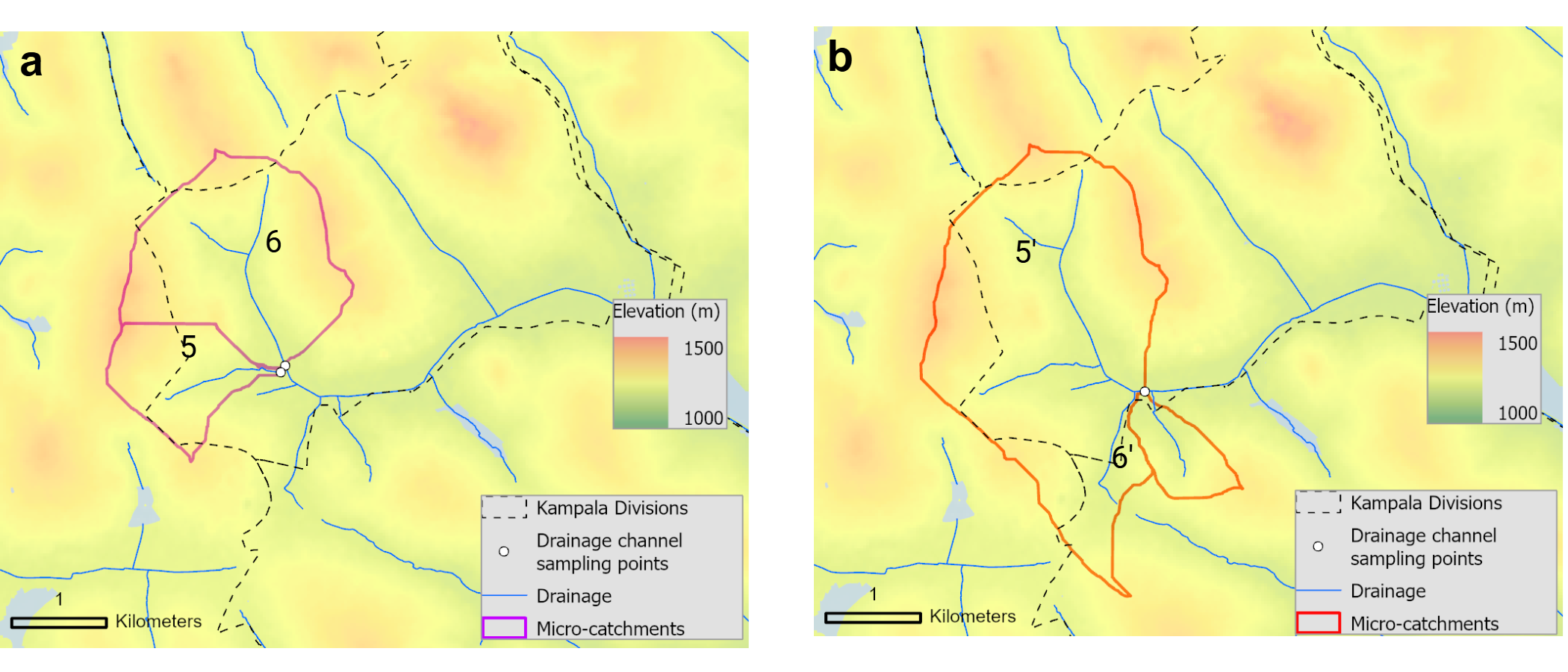

**Fig. S1** Micro-catchments from two drainage channel sampling points, #5 and 6, during two sampling campaigns in March (a) and September/October (b). Due to drainage channel construction, the original sampling points in March were blocked in September. The alternative two points downstream 0.6 km are annotated as #5’ and 6’.

### Text S1 Measurement of volumetric flow rate of drainage channels

At the time of sampling, flow velocity (m/s) was measured using a portable flowmeter (FLO-MATE^TM^, Marsh-McBirney, Inc.) in the first campaign and timing the travel of a floating object over a predetermined distance of ~200 cm in the second campaign. Additionally, the depth (m) and width (m) of each water flow in the channel were measured. These parameters were used to calculate the volumetric flow rate (m^3^/day) for each sampling point.

### Text S2 Sample concentration and DNA/RNA extraction

Upon arrival of samples at the laboratory at Makerere University, each grab sample was aliquoted to 50 mL, treated with MgCl_2_ to achieve a final concentration of 25 mM. Then, samples were vacuum-filtered using S-Pak® membrane filters (mixed cellulose esters, pore size 0.45 μm, diameter 47 mm; Merck, Cat. No. HAWG047S6). The DNA/RNA was extracted from the filters using AllPrep PowerFecal Pro DNA/RNA Kit (Qiagen, Cat. No. 80254) which were subsequently torn into small pieces with sterile forceps and transferred into the PowerBead Pro Tube and then processed according to the manufacturer's protocol. DNA and RNA were each eluted in 100 µL of DNAse/RNAse-free water and collected separately following the protocol of the AllPrep kit, yielding two tubes of 100 µL each. To remove potential PCR inhibitors, the 100 μL extracted DNA and RNA were further purified using the Zymo OneStep PCR inhibitor removal kit (Zymo Research, Cat. No. D6030). There were no extraction replicates. Extraction blanks were included with each day of sampling, prepared using 50 mL of nuclease-free water and processed in parallel following the same procedure. Purified extracts were 3× diluted with nuclease-free water to minimize potential environmental inhibition and stored at −80 °C until being transferred on dry ice to the laboratory at the Eawag in Dübendorf, Switzerland. Upon arrival, extracts were stored at −80 °C, for ≤12 weeks for the first campaign and ≤ 21 weeks for the second campaigns, before analysis.

**Table S1** Sequence of digital PCR primers and probes used in this study. Those probes contained fluorescent molecules (HEX, FAM, Cy5, ROX, ATTO425) and quenchers (BHQ-1, BHQ-2, MGBQ530).

|  | Target | Sequences (5’-3’) | | Reference |
| --- | --- | --- | --- | --- |
| Duplex | Rotavirus | Forward | GGCTTTTAAAGCGTCTCAGT | ^1^ |
|  |  | Reverse | AATYTATAGCTATCRTTCTCYARATG |  |
|  |  | Probe | HEX/CCATGGCTGAGCTAGCTTGCTT/MGBQ530 |  |
|  | Norovirus GII | Forward | ATGTTYAGRTGGATGAGATTCTC | ^2^ |
|  |  | Reverse | TCGACGCCATCTTCATTCAC |  |
|  |  | Probe | Cy5/TGAGCACGTGGGAGGGCGATCGC/BHQ-2 |  |
| Singleplex | Norovirus GI | Forward | CGYTGGATGCGNTTYCATGA | ^2^ |
|  |  | Reverse | CTTAGACGCCATCATCATTYAC |  |
|  |  | Probe | FAM/TGGACAGGRGATCGCRATCT/MGBQ530 |  |
|  | PMMoV | Forward | GAGTGGTTTGACCTTAACGTT | ^3^ |
| Singleplex |  | Reverse | TTGTCGGTTGCAATGCAAGT |  |
|  |  | Probe | BHQ-1/CCTACCGAAGCAAATG/FAM |  |
| Six-plex | SARS-CoV-N1 | Forward | GACCCCAAAATCAGCGAAAT | ^4^ |
|  |  | Reverse | TCTGGTTACTGCCAGTTGAATCTG |  |
|  |  | Probe | ATTO425/ACCCCGCATTACGTTTGGTGGACC/BHQ-1 |  |
|  | SARS-CoV-N2 | CDC Kit, 2019-nCoV RUO Kit (Integrated DNA Technologies),  FAM-labelled probe | |  |
|  | Influenza A virus | Forward | TGGAATGGCTAAAGACAAGACCAAT | ^5^ |
|  |  | Reverse | AAAGCGTCTACGCTGCAGTCC |  |
|  |  | Probe | Cy5/TTTGTKTTCACGCTCACCGTGCCC/BHQ-2 |  |
|  | Influenza B virus | Forward | GAGACACAATTGCCTACYTGCTT | ^5^ |
|  |  | Reverse | ATTCTTTCCCACCRAACCAACA |  |
|  |  | Probe | HEX/AGAAGATGGAGAAGGCAAAGCAGAACTAGC/BHQ-1 |  |
|  | Respiratory Syncytial Virus | Forward | CTCCAGAATAYAGGCATGAYTCTCC | ^6^ |
|  |  | Reverse | GCYCTYCTAATYACWGCTGTAAGAC |  |
|  |  | Probe | ROX/TAACCAAATTAGCAGCAGGAGATAGATCAG/BHQ-2 |  |
| Triplex | *V. cholerae*-*ompW* | Forward | CACCAAGAAGGTGACTTTATTGTG | ^7^ |
|  |  | Reverse | GGAAAGTCGAATTAGCTTCACCAA | Self-designed |
|  |  | Probe | FAM/ACATAAGATTTCTACCTCTGGTGGT/BHQ-1 | Self-designed |
|  | *V.cholerae*-*O1rfb* | Forward | GTTTCACTGAACAGATGGG | ^8^ |
|  |  | Reverse | GGTCATCTGTAAGTACAAC | ^8^ |
|  |  | Probe | HEX/CATGCCTATTCTGACGTAAT/BHQ-1 | Self-designed |
|  | *V. cholerae*-*O139rfb* | Forward | TGGGATGCCAGTCATGCTGT | Self-designed |
|  |  | Reverse | GTCAAACCCGATCGTAAAGG | ^8^ |
|  |  | Probe | Cy5/CACTGTGGTGGGTATTTTAC/BHQ-2 | Self-designed |

### Text S3 Digital PCR assays for pathogen quantification

Multiple pathogens were quantified using a one-step digital RT-PCR or PCR assays on the naica® PCR platform (Stilla Technologies, Villejuif, France). Pepper Mild Mottle Virus (PMMoV) was quantified as a fecal indicator and a concentration normalizer due to its high prevalence and stability in human feces.^9^ Norovirus GI was quantified using a singleplex assay. Norovirus GII and Rotavirus were quantified using a duplex assay. Respiratory viruses were measured using a six-plex assay^10^ targeting SARS-CoV-2 (N1 and N2 regions), Influenza A and B viruses, Respiratory syncytial virus (RSV), and Murine Hepatitis Virus (MHV). MHV, used as a spike-in control in the Swiss wastewater surveillance program^11^, was not targeted in this study. A triplex PCR assay was used for quantification of *Vibrio cholerae* (*V. cholerae*), targeting the species-specific gene (*ompW*) and the serogroup markers (*O1rfb* and *O139rfb*).

All primers and probes (**Table S1**) were purchased from Microsynth AG (Balgach, Switzerland) except for the SARS-CoV-2 CDC Kit from Integrated DNA Technologies (IDT; Coralville, USA, Cat. No. 10006713) and shipped in solid-state. Upon arrival, they were reconstituted to 100 µM in nuclease-free water and stored at −20 °C. Mastermix preparation and template addition were conducted in separate PCR workspaces to minimize cross-contamination. For RNA targets, each reaction contained 13.5 μL of 2× qScript XLT One-Step RT-qPCR ToughMix (Quantabio, Beverly, MA, USA, Cat. No. 95132), 0.1 μM fluorescein sodium salt (0.2 μM for the PMMoV assay; VWR International GmbH, Dietlikon, Switzerland, Cat. No. 0681-100G), 0.5 μM forward and reverse primers, 0.2 μM probes. Nuclease-free water was added to reach 21.6 µL, followed by 5.4 µL of RNA template. For Norovirus GII and Rotavirus duplex assay, the RNA templates were denatured by incubating at 95 °C for 5 min than cooling on ice to heat snap the Rotavirus.^12^ For DNA targets, each reaction contained 2.7 and 1.08 µL of Buffer A and B from the naica® multiplex PCR Mix (Stilla Technologies, Villejuif, France; Cat. No. R10104) and 0.1 µM of fluorescein sodium salt, according to manufacturer's recommendations, along with 0.5 μM forward and reverse primers, and 0.2 μM probes. Nuclease-free water was added to reach a volume of 21.6 µL, followed by 5.4 µL of DNA template for a prereaction volume of 27 µL.

After adding the template, the reaction mixture was gently vortexed and spun down and a reaction volume of 25 μL was loaded on Sapphire chips (Stilla Technologies, Villejuif, France; Cat. No. C14012). All reactions were performed in duplicate with one positive control and one no-template control (NTC) for every five samples processed. The chips were processed using a naica® Geode (Stilla Technologies, Villejuif, France) system, which partitions the reaction mixture into droplets (approximately 0.519 nL per droplet) for 12 min at 40 °C before thermocycling. For PMMoV, thermocycling consisted of reverse transcription at 50 °C for 1 h, enzyme activation at 95 °C for 5 min, and 45 cycles of 95 °C for 15 s and 60 °C for 1 min. For Norovirus GI (singleplex) and Rotavirus/Norovirus GII (duplex) assays, reverse transcription was performed at 50 °C for 1 h, enzyme activation at 95 °C for 5 min, and 45 cycles of 95 °C for 15 s and 54 °C for 1 min. For the respiratory six-plex assay, thermocycling consisted of reverse transcription at 50 °C for 1 h, enzyme activation at 95 °C for 5 min, 40 cycles of 95 °C for 30 s and 57.5 °C for 1 min. For the triplex *V. cholerae* assay, the reaction condition consisted of 45 cycles of 95 $^{\circ}$C for 10 s and 59 $^{\circ}$C for 1 min.

After thermocycling, the release step that lowers the pressure and temperature was followed and the chips were transferred to the naica® Prism3 (or Prism6 for respiratory six-plex assay) system (Stilla Technologies, Villejuif, France) for fluorescence imaging. Droplets were analyzed using Crystal Miner software (version 4.0, Stilla Technologies, Villejuif, France), with the minimum number of 15,000 analyzable required for quality control. If one duplicate did not meet the minimum number, the sample was repeated. Positive and negative droplets were classified based on visual thresholding of fluorescence signals across multi-color channels, and absolute concentrations (C_PCR,_ gc/µL) were calculated using Poisson distribution analysis. The sample was considered detectable if the number of positive droplets per well was three or more, which corresponds to ≥ 0.23 gc/µL reaction volume, or ≥ 8.1 gc/mL sample volume, for 20,000 analyzable droplets. Extraction blanks were included to assay cross contamination during sample collection and processing. A limit of blank (LOB) of 5 positive droplets was applied as a quality control criterion across all assays. Extraction blanks were five positive droplets or below for all targets, with exception of one Rotavirus extraction blank with exceed the LOB with 8 positive droplets; all corresponding Rotavirus data from that sampling day were excluded from further analysis. Full extract blank results for all assays are provided in **Table S3**.

Sample concentrations (C_sample,_ gc/mL) were determined based on dilution factors (D.F.) and volume ratios between reaction volume (*V_rxn_*), template input (*V_template_*), extraction elution volume (*V_elution_*), and initial sample volume (*V_sample_*) using the following equation:

C_sample_ $\left( \frac{gc}{mL} \right)$ = C_PCR_ $\left( \frac{gc}{\mu L} \right)$ $\times$ D. F. $\times$ $\frac{V_{rxn} (\mu L)}{V_{template} (\mu L)} \times\frac{V_{elution} (\mu L)}{V_{sample} (mL)}$

**Table S2** The number of positive droplets by dPCR assays in the blank extracts

| # of positive droplets | SARS-N1 | SARS-N2 | Influenza A | Influenza B | RSV | PMMoV | Rotavirus | Norovirus GI | Norovirus GII | ompW | O1rfb | O139rfb |
| --- | --- | --- | --- | --- | --- | --- | --- | --- | --- | --- | --- | --- |
| 17-Mar | 2 | 0 | 0 | 0 | 0 | 0 | 0 | 1 | 0 | 0 | 0 | 0 |
| 18-Mar | 0 | 0 | 1 | 0 | 0 | 0 | 1 | 0 | 0 | 0 | 0 | 0 |
| 19-Mar | 1 | 0 | 0 | 0 | 0 | 0 | 0 | 0 | 0 | 1 | 0 | 0 |
| 20-Mar | 0 | 0 | 0 | 0 | 0 | 0 | 0 | 0 | 0 | 0 | 0 | 0 |
| 21-Mar | 0 | 0 | 0 | 0 | 0 | 0 | 5 | 2 | 0 | 3 | 0 | 0 |
| 24-Mar | 1 | 0 | 0 | 0 | 0 | 0 | 3 | 0 | 0 | 0 | 0 | 0 |
| 25-Mar | 0 | 0 | 0 | 0 | 0 | 1 | 3 | 1 | 0 | 0 | 0 | 0 |
| 26-Mar | 0 | 0 | 0 | 0 | 0 | 1 | 0 | 0 | 0 | 0 | 0 | 0 |
| 27-Mar | 0 | 0 | 0 | 0 | 0 | 0 | 0 | 0 | 0 | 0 | 0 | 0 |
| 29-Mar | 0 | 0 | 0 | 0 | 0 | 0 | 0 | 0 | 0 | 0 | 0 | 0 |
| 15-Sep | 0 | 0 | 0 | 0 | 0 | 0 | 0 | 0 | 0 | 0 | 0 | 0 |
| 16-Sep | 0 | 0 | 0 | 0 | 0 | 0 | 0 | 0 | 0 | 0 | 0 | 0 |
| 18-Sep | 1 | 0 | 0 | 0 | 0 | 0 | 0 | 0 | 0 | 0 | 0 | 0 |
| 19-Sep | 0 | 0 | 0 | 0 | 0 | 1 | 0 | 0 | 0 | 2 | 0 | 0 |
| 22-Sep | 0 | 0 | 0 | 0 | 0 | 1 | 8* | 2 | 0 | 0 | 0 | 0 |
| 23-Sep | 0 | 0 | 0 | 0 | 0 | 0 | 0 | 0 | 0 | 0 | 0 | 0 |
| 25-Sep | 0 | 0 | 0 | 2 | 2 | 0 | 1 | 0 | 0 | 0 | 0 | 0 |
| 26-Sep | 0 | 0 | 0 | 0 | 0 | 0 | 1 | 0 | 0 | 0 | 0 | 0 |
| 27-Sep | 0 | 0 | 0 | 0 | 0 | 0 | 3 | 0 | 0 | 0 | 0 | 0 |
| 29-Sep | 0 | 0 | 0 | 0 | 0 | 0 | 0 | 0 | 0 | 0 | 0 | 0 |
| 30-Sep | 0 | 0 | 0 | 0 | 0 | 0 | 0 | 0 | 0 | 0 | 0 | 0 |
| 2-Oct | 0 | 0 | 0 | 0 | 0 | 3 | 3 | 0 | 0 | 0 | 0 | 0 |
| 3Oct | 0 | 0 | 1 | 2 | 2 | 0 | 0 | 0 | 0 | 1 | 0 | 0 |
| 6-Oct | 0 | 0 | 0 | 0 | 0 | 0 | 0 | 0 | 0 | 1 | 0 | 0 |
| 7-Oct | 0 | 0 | 0 | 0 | 0 | 0 | 0 | 0 | 0 | 1 | 0 | 0 |
| 8-Oct | 0 | 0 | 0 | 0 | 0 | 0 | 5 | 0 | 0 | 1 | 0 | 0 |
| 10-Oct | 0 | 0 | 0 | 0 | 0 | 0 | 0 | 0 | 0 | 1 | 0 | 0 |

*exceeds the limit of blank (LOB), six or more positive droplets

**Table S3** Inhibition test against SARS-CoV-2 N1 on the subset of drainage channel and WWTP influent samples

| Inhibition (%) | | SARS-CoV-2 N1 |
| --- | --- | --- |
| 28-Mar | #1 | -3 |
|  | #2 | -11 |
|  | #3 | -18 |
|  | #4 | -10 |
|  | #5 | 6 |
|  | #6 | -21 |
|  | #7 | -1 |
|  | #8 | -6 |
|  | #9 | -7 |
|  | #10 | -9 |
|  | Lubigi | -10 |
|  | Nakivubo | -6 |
| 10-Oct | #1 | -3 |
|  | #2 | -13 |
|  | #3 | -10 |
|  | #4 | -6 |
|  | #5’ | 0 |
|  | #6’ | -19 |
|  | #7 | -14 |
|  | #8 | -18 |
|  | #9 | -9 |
|  | #10 | -2 |
|  | Lubigi | -10 |
|  | Nakivubo | -10 |

PCR inhibition was evaluated on a subset of samples (n=20 for drainage channel samples, n=4 for WWTP influents) using digital PCR assay for SARS-CoV-2 targeting N1 region.^10^ Apart from the concentration of SARS-CoV-2 in the original sample (C_original_). A known quantity of SARS-CoV-2 RNA (C_spike_) was spiked into the extract and the total SARS-N1 concentration was measured (C_observed_).

Inhibition was calculated using the following equation:

Inhibition (%) = 1 − (C_observed_/ (C_spike_+ C_original_))

An inhibition value of 0% indicated no inhibition.

### Text S4 *E. coli* enumeration and physicochemical analysis

Phosphate-buffered saline (PBS) (10×, pH 7.4) (Thermo-Fisher, Zürich, Switzerland; Cat. No. AM9625) was used to serially dilute the samples. Serial dilution by 10-fold from 10^-2^ to 10^-4^ was conducted, and 1 mL of the samples was pipetted in the CompactDry® EC plate (Shimadzu Diagnostics Corporation, Tokyo, Japan; Cat. No. MSDS54052) and stored at 37 °C for 24 hours. Then, the colony-forming unit (CFU) of *E. coli* was estimated by counting blue/blue-purple-colored colonies, according to manufacturer's instructions.

Total suspended solids (TSS) (mg/L) were measured for all collected WWTP influents and drain samples using S-Pak® Membrane Filter (mixed cellulose esters (MCE), pore size 0.45 μm, filter diam. 47 mm, Merck, Cat. No. HAWG047S6). The oven-dried crucible and filter were prepared at 105 °C for 1 hour and weighed after cooling at room temperature in a desiccator. Five to 100 mL of drain or WWTP influent samples that yield between 2.5 and 200 mg residue were filtered and dried at 105 °C for 24 hours in the crucible. TSS was calculated as the difference between the dry filter mass after filtration and the initial filter mass before filtration, divided by the filtered volume.

**Table S4** Meteorological conditions during the two sampling campaigns in Kampala, Uganda

|  | March  2025 | September 2025 | October  2025 | Sep/Oct  2025 |
| --- | --- | --- | --- | --- |
| Sampling durations (days) | 12 | 16 | 10 | 26 |
| Monthly total rainfall (mm) | 213·9 | 44.1 | 154.0 | 198·1 |
| *Rainy days in month (n) | 13 | 7 | 14 | 21 |
| Rainfall during sampling period (mm) | 148·1 | 36.2 | 66.5 | 102·7 |
| Sampling period (day) | 12 |  |  | 26 |
| Average rainfall during the sampling period (mm/day) | 12·3 |  |  | 4·0 |

*Rainy days defined as days with more than 1 mm of rainfall

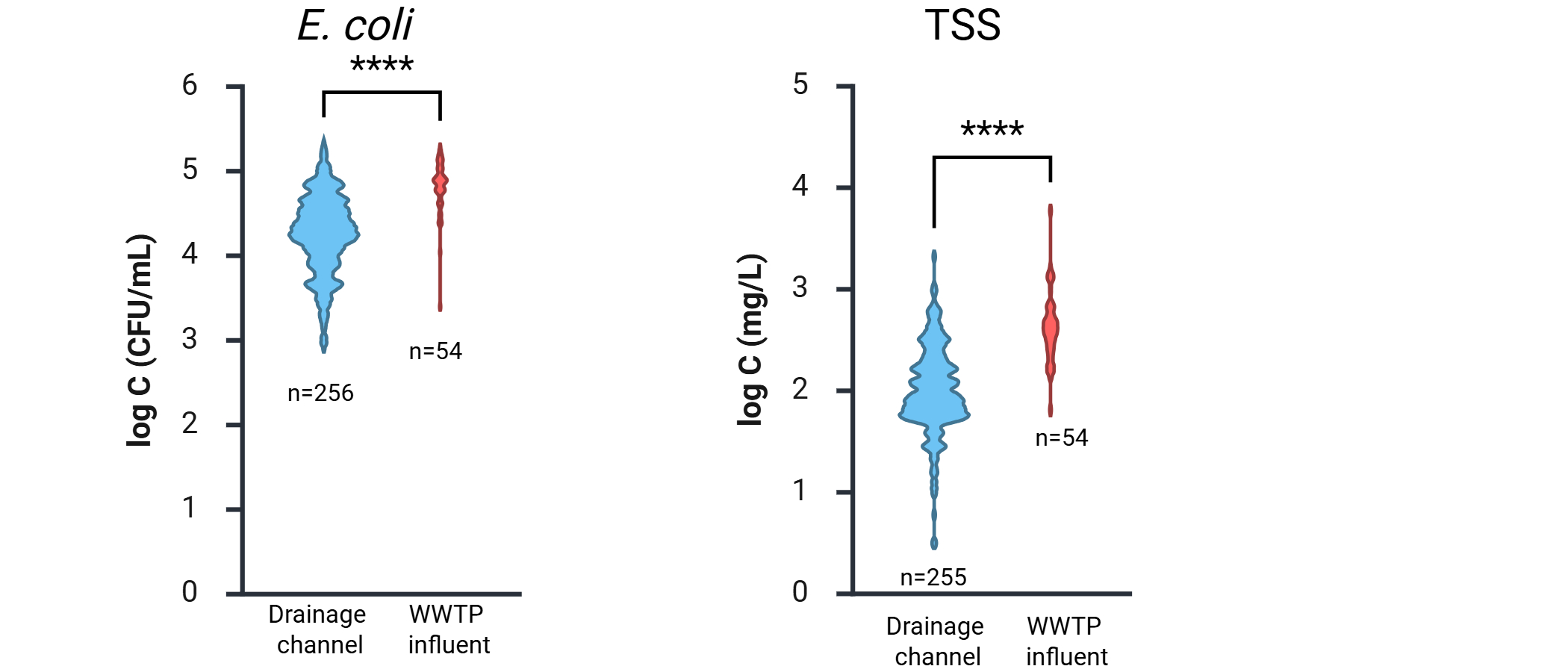

**Fig. S2** Violin plots of log_10_ concentrations of *E. coli* and Total suspended solids (TSS) across ten drainage channel sampling points and two WWTP influents. The number of samples and statistical difference between drains and WWTP influents are indicated (Two-way ANOVA with Bonferroni, **** p<0·0001).

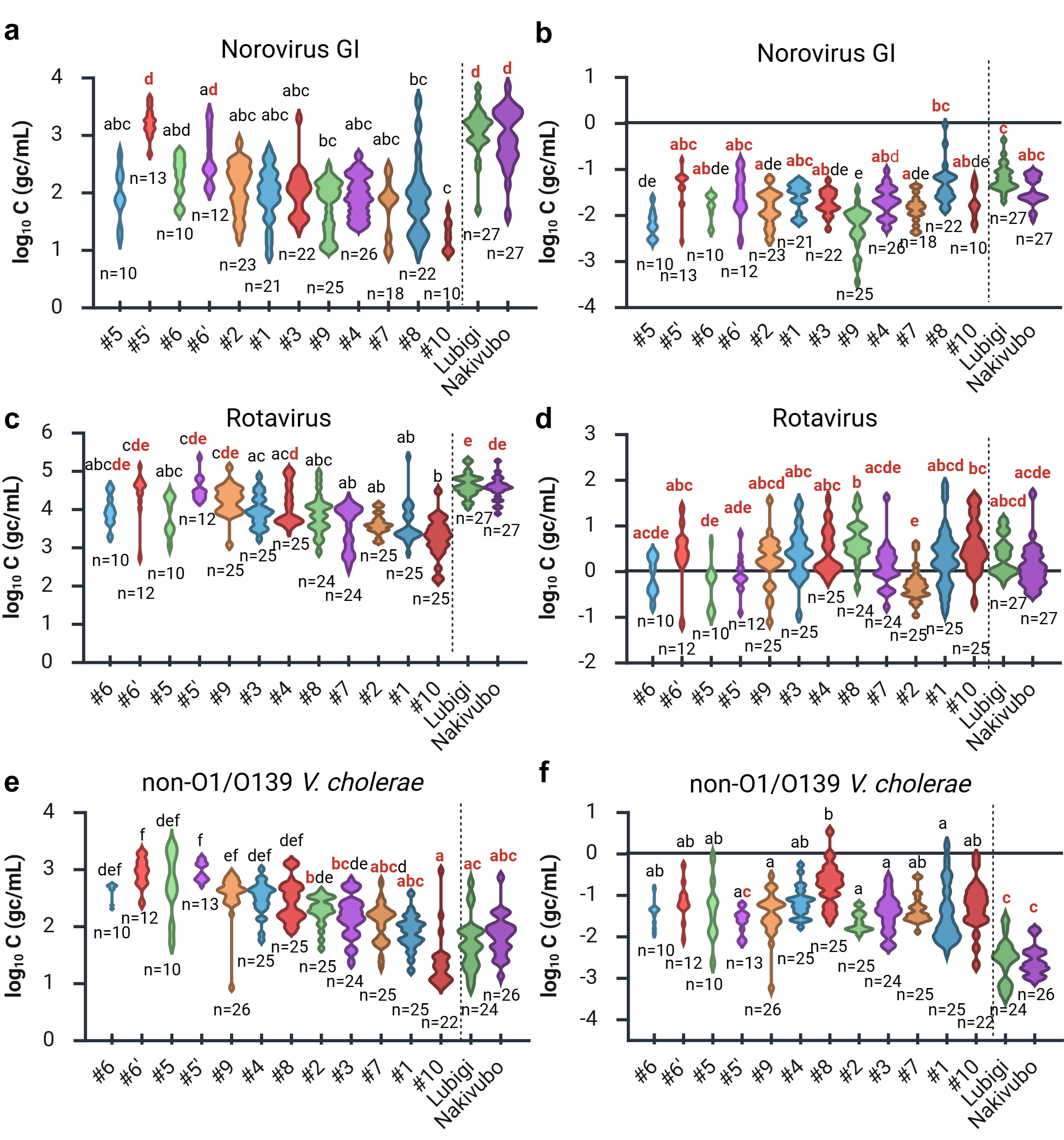

**Fig. S3** Violin plots of log_10_ concentrations of (a, c, d) Norovirus GI, Rotavirus, and non-O1/O139 *V. cholerae* and (b, d, f) their PMMoV-normalized concentrations across ten drainage channel sampling points, #1-10, and two WWTP influents, Lubigi and Nakivubo, ordered from left to right by ascending median concentration for drainage channels, with two WWTP influents on the right. Violin plots are scaled by count with light smoothing. Group differences are summarized using compact letter display (CLD) based on Kruskal-Wallis test with Dunn's pairwise multiple comparisons; CLD letters shared between WWTP influents and drainage channels are highlighted in red.

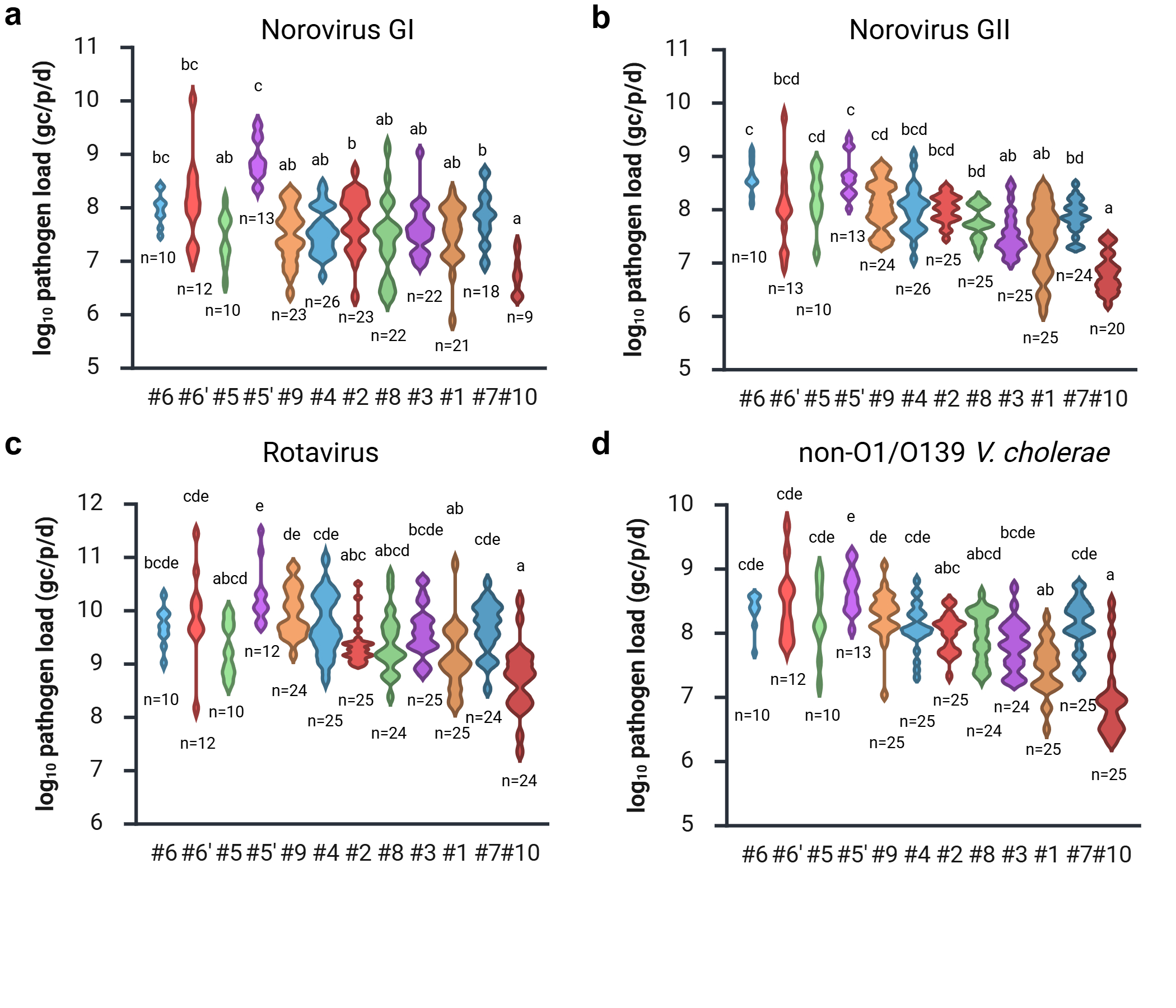

**Fig. S4** Violin plots of log_10_ daily pathogen load normalized by micro-catchment population and flow rate (gc/population/day) of Norovirus GI and GII, Rotavirus, and non-O1/O139 *V. cholerae* across ten drainage channel sampling points, #1-10, ordered from left to right by ascending median absolute concentration for drainage channels. Violin plots are scaled by count with light smoothing. Group differences are summarized using compact letter display (CLD) based on Kruskal-Wallis test with Dunn's pairwise multiple comparisons.

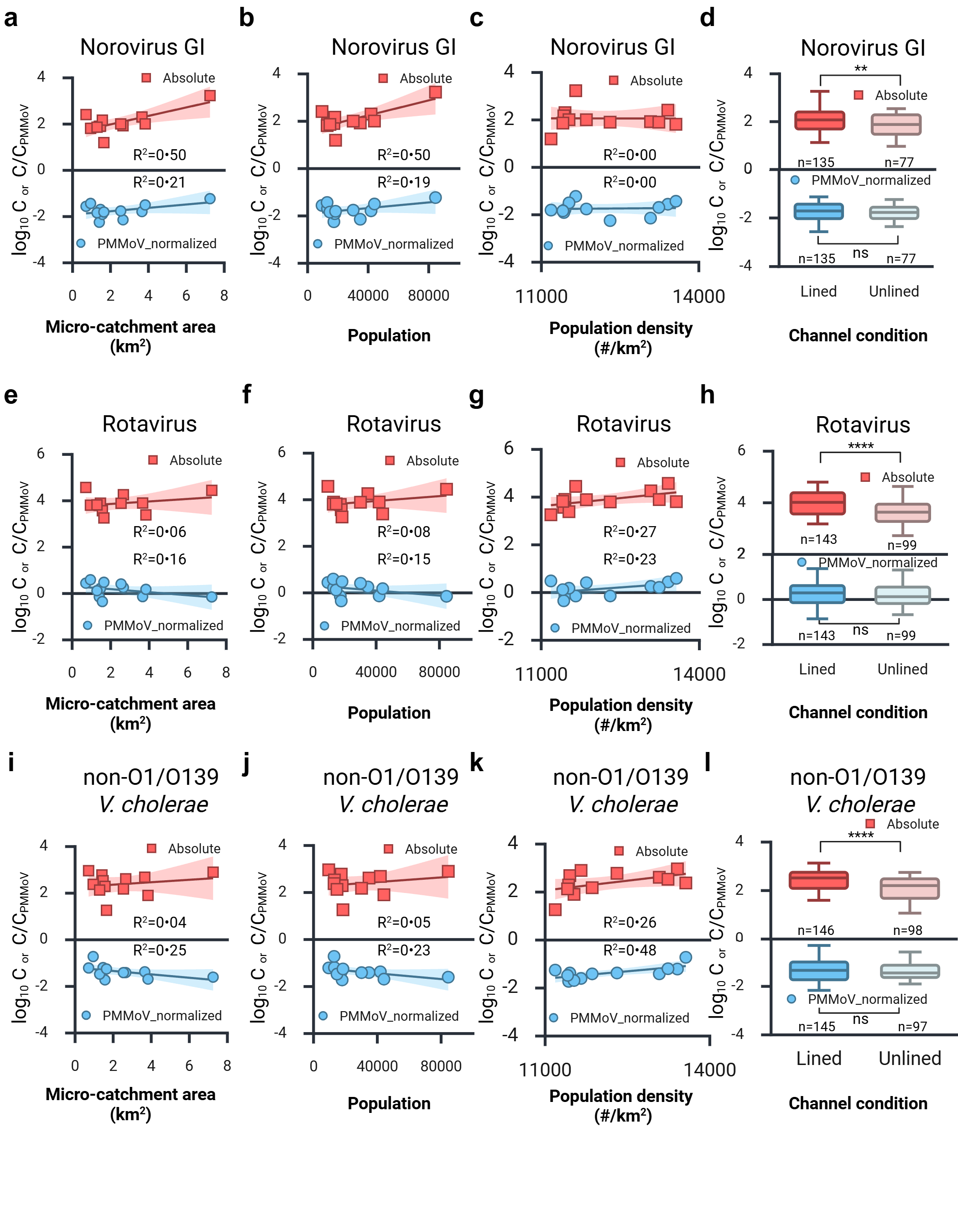

**Fig. S5** **Association between concentrations and micro-catchment characteristics.** Scatter plots of median log_10_ concentrations (upper panels) and PMMoV-normalized concentrations (lower panels) of Norovirus GI (a-c), Rotavirus (e-g), and non-O1/O139 *V. cholerae* (i-k) across micro-catchments by catchment area (km^2^), population, and population density (#/km^2^). The line and shaded region indicate best-fit linear regression with 95% confidence interval and R-square value. (d, h, l) Boxplot of log_10_ concentrations (upper) and PMMoV-normalized concentrations (lower) of Norovirus GI (d), Rotavirus (h), and non-O1/O139 *V. cholerae* (l) by drainage channel condition (lined vs. unlined), showing median and 5^th^-95^th^ percentile range. Mann-Whitney U test was used for (g). (**** p<0·0001; (** p<0·01; ns: not significant)

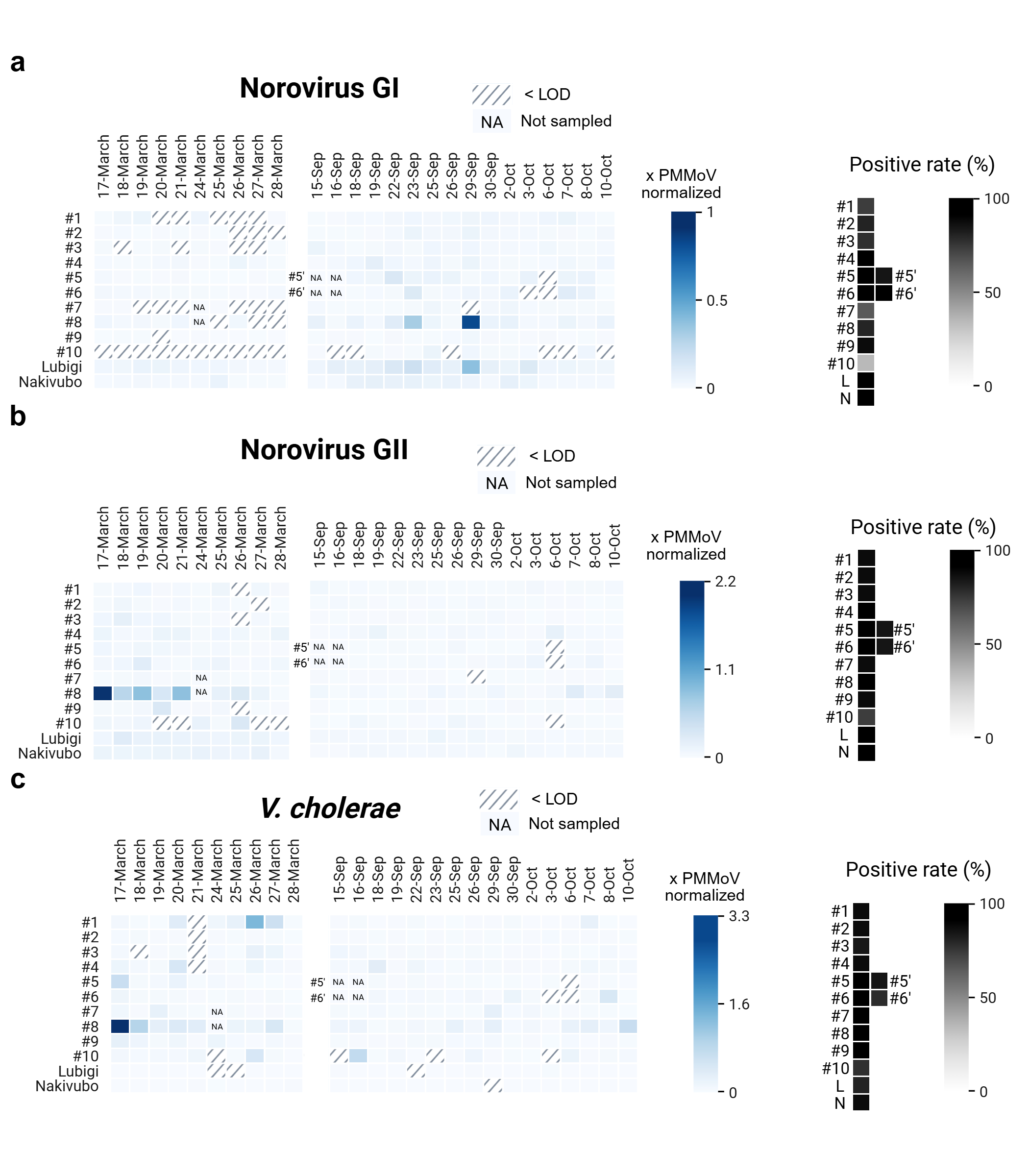

**Fig. S6** Temporal dynamics of PMMoV-normalized pathogen concentrations in drainage channels and wastewater influents to the centralized treatment plants (WWTPs) Heatmaps of PMMoV-normalized concentrations of Norovirus GI (a), Norovirus GII (b), and non-O1/O139 *Vibrio cholerae* (*V. cholerae*) (c) across ten drainage channel sampling points, #1-10, and two WWTP influents, Lubigi and Nakivubo, over two sampling campaigns from 17 to 28 March 2025 and from 15 September to 10 October 2025. Hatched cells indicate concentrations below the limit of detection (LOD); “NA” indicates missing samples. On each right panel, the heat maps of positive rate of samples ($\%$) are shown for individual drainage channel sampling point and each WWTP influent, annotated L (Lubigi) and N (Nakivubo).

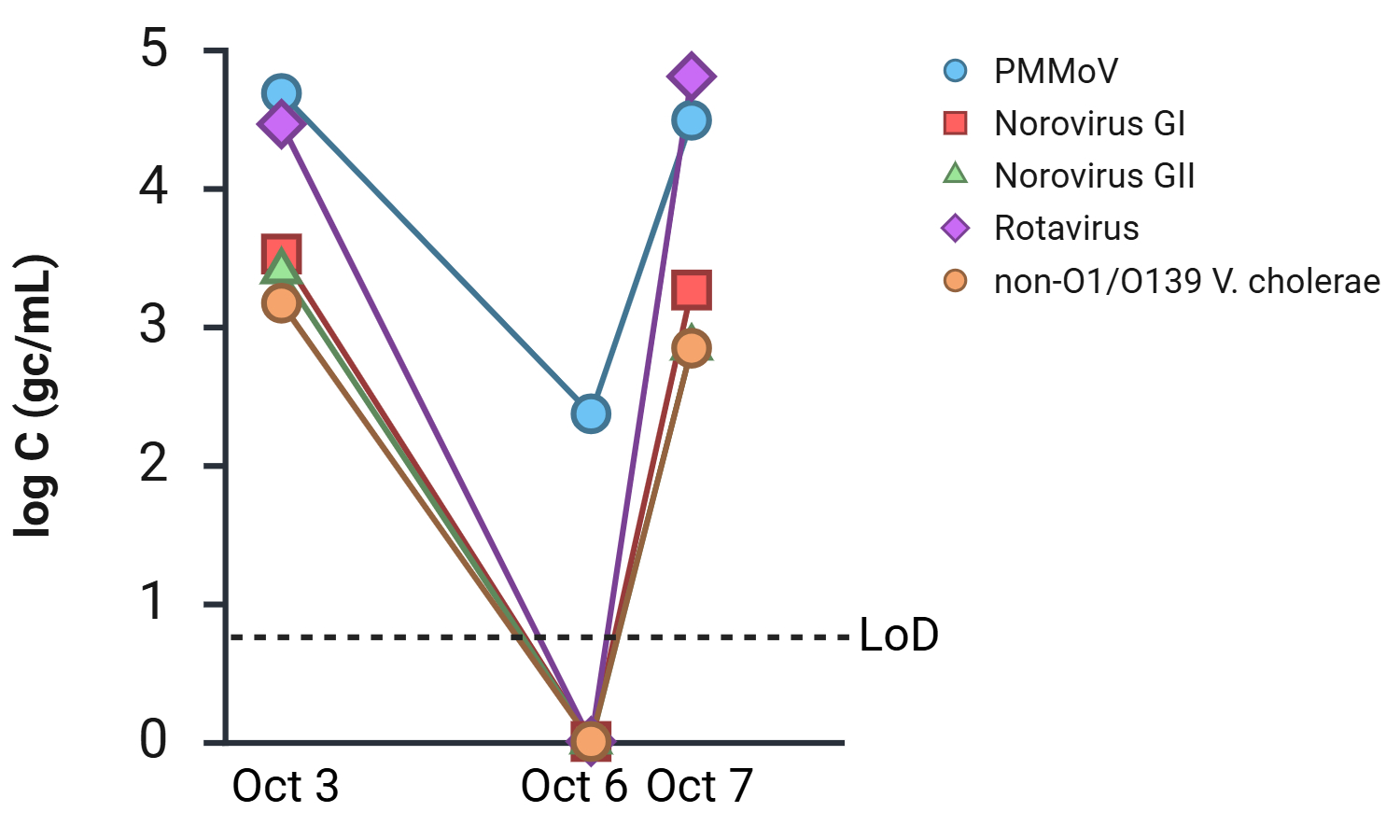

**Fig. S7** Impact of a flooding event on pathogen concentrations at drainage channel sampling point, #5’. Concentrations of PMMoV, Norovirus GI, Norovirus GII, Rotavirus, and non-O1/O139 *V. cholerae* (log gc/mL) are shown for October 3, 6, and 7. There was a flooding event at #5’ on Octboer 6.

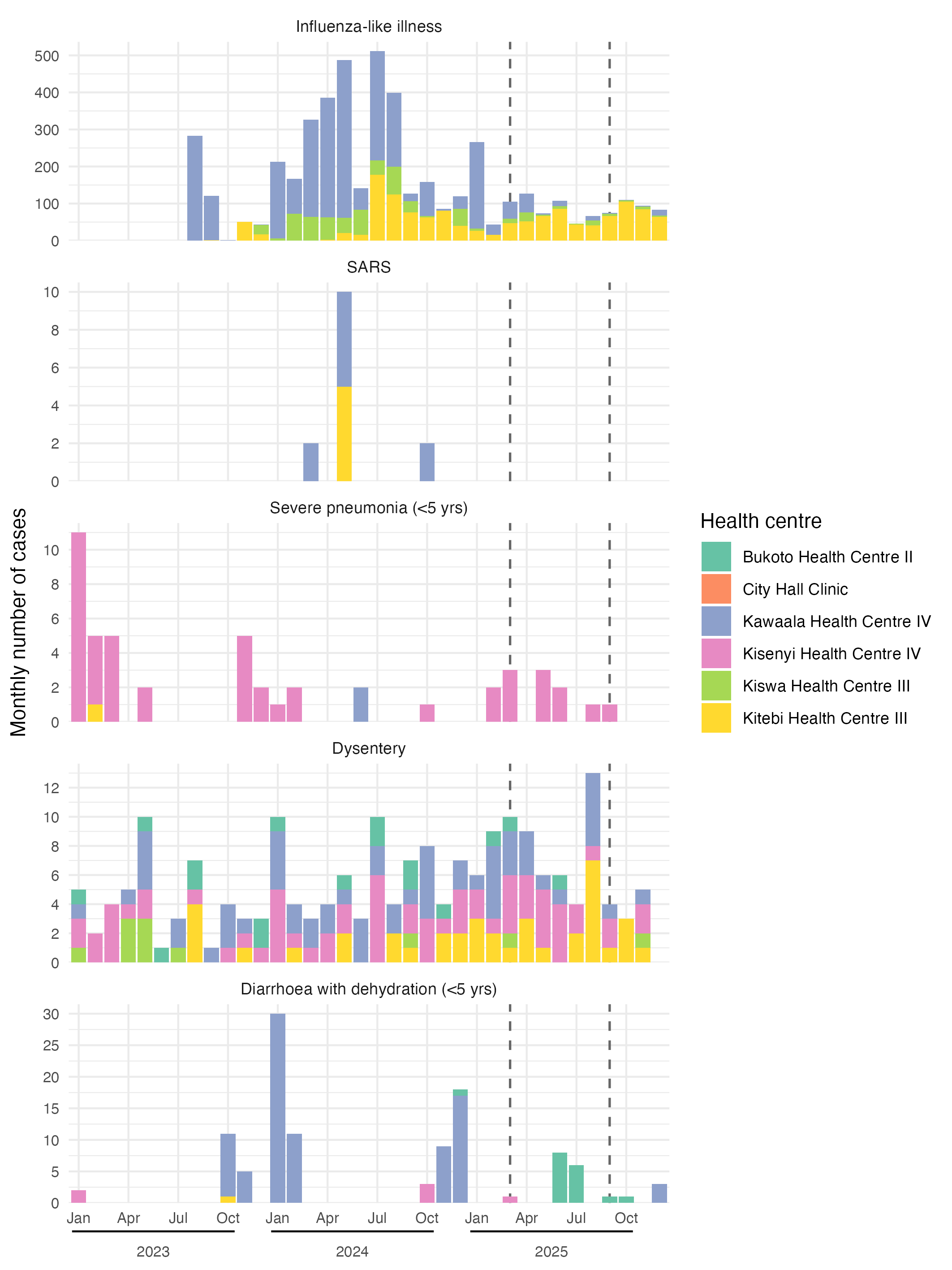

**Fig. S8** Monthly number of respiratory and diarrheal disease cases recorded through routine surveillance at official health centers located near sampling points. The vertical dotted lines indicate the start of the two sampling rounds (March 2025 and September 2025).

**Table S5** Annual number of respiratory and diarrheal disease cases recorded through routine surveillance at official health centers located near sampling points.

| **Year** | **Health centre** | **Disease (number of reported cases)** | | | | |
| --- | --- | --- | --- | --- | --- | --- |
|  |  | Influenza-like illness | SARS | Severe pneumonia  (<5 yrs) | Dysentery | Diarrhea with dehydration (<5 yrs) |
| 2023 | Bukoto Health Centre II | 0 | 0 | 0 | 7 | 0 |
|  | City Hall Clinic | 0 | 0 | 0 | 0 | 0 |
|  | Kawaala Health Centre IV | 405 | 0 | 0 | 13 | 15 |
|  | Kisenyi Health Centre IV | 0 | 0 | 29 | 15 | 2 |
|  | Kiswa Health Centre III | 25 | 0 | 0 | 8 | 0 |
|  | Kitebi Health Centre III | 69 | 0 | 1 | 5 | 1 |
| 2024 | Bukoto Health Centre II | 0 | 0 | 0 | 7 | 1 |
|  | City Hall Clinic | 0 | 0 | 0 | 0 | 0 |
|  | Kawaala Health Centre IV | 2019 | 9 | 2 | 26 | 67 |
|  | Kisenyi Health Centre IV | 0 | 0 | 4 | 26 | 3 |
|  | Kiswa Health Centre III | 503 | 0 | 0 | 1 | 0 |
|  | Kitebi Health Centre III | 601 | 5 | 0 | 10 | 0 |
| 2025 | Bukoto Health Centre II | 0 | 0 | 0 | 3 | 16 |
|  | City Hall Clinic | 0 | 0 | 0 | 0 | 0 |
|  | Kawaala Health Centre IV | 410 | 0 | 0 | 21 | 3 |
|  | Kisenyi Health Centre IV | 0 | 0 | 12 | 25 | 1 |
|  | Kiswa Health Centre III | 91 | 0 | 0 | 2 | 0 |
|  | Kitebi Health Centre III | 696 | 0 | 0 | 24 | 0 |
| **Total** |  | 4819 | 14 | 48 | 193 | 109 |
