## Supplementary material for "Wastewater and Environmental Surveillance in Cities with Sewered and Non-sewered Sanitation: Evidence from Kampala, Uganda": Annex 1. Group discussion and field validation

Focus Group Discussion – Expert Group Interview

First focus group discussions were held on March 12, 2025, with the experts at KCCA and Eawag, followed by two days of field validation for 22 sampling points on March 12 and 13, 2025.

Then, another group discussion on March 15, 2025, with an expert at Makerere University who involved the field validation to narrow down 10 sampling points for experiment.

Last group discussion was conducted on April 1, 2025, involving the same experts at KCCA and Makerere University, to re-validate our assessment and discussion on 10 sampling points.

CAUTION: The annotations of selected 10 sampling points (#1, 8, 9, 10, 13, 14, 16, 19, 21, 22) were recorded as #1 to #10 in the Manuscript.

| Group discussion report | Manuscript |
| --- | --- |
| #1 | #1 |
| #8 | #2 |
| #9 | #3 |
| #10 | #4 |
| #13 | #5 |
| #14 | #6 |
| #16 | #7 |
| #19 | #8 |
| #21 | #9 |
| #22 | #10 |

**Group discussion on March 12, 2025**

**Experts at Kampala Capital City Authority (KCCA) and Eawag**

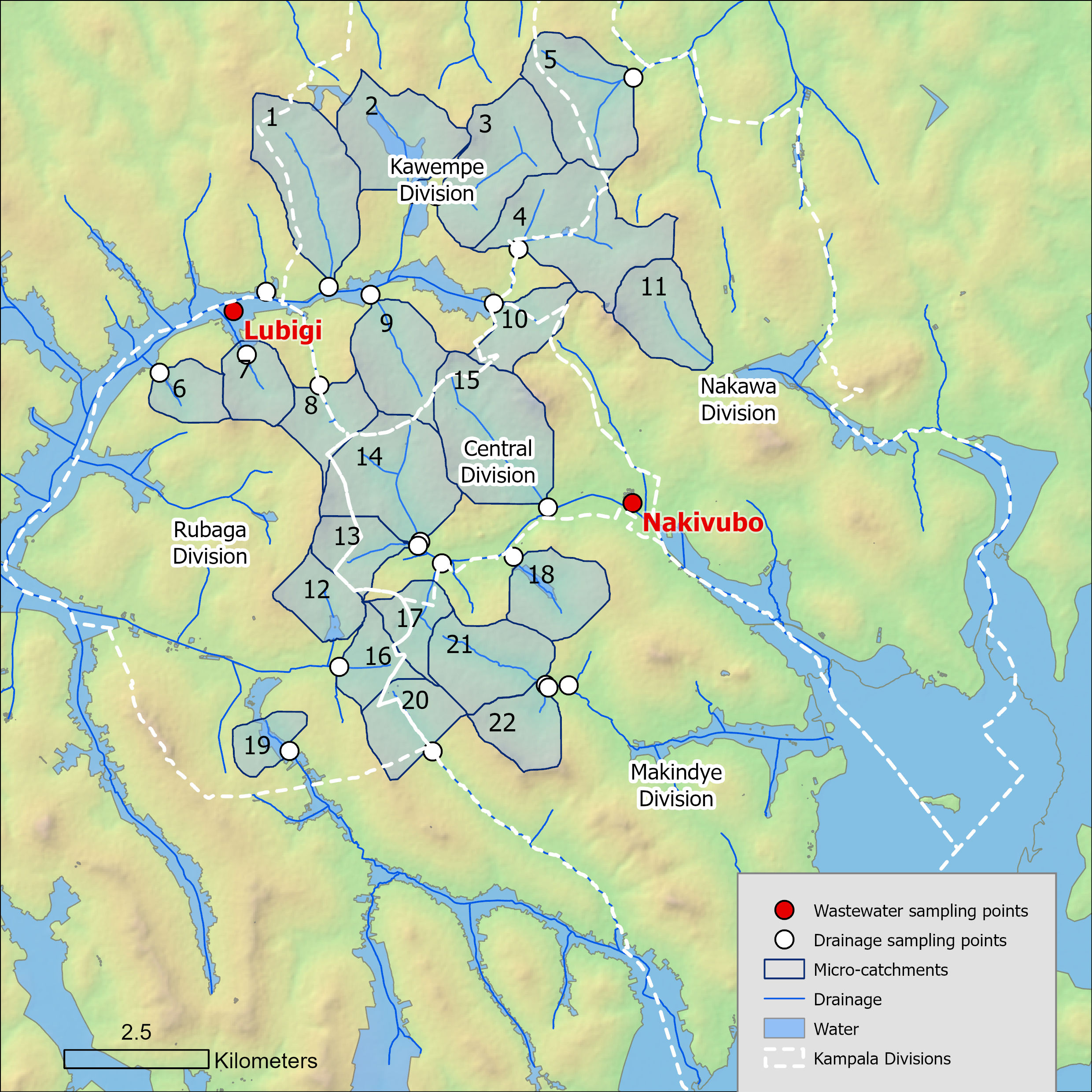

Sampling points for 22 micro-catchments were explained, based on maps of elevation and surface water, and trying to get a diversity of locations based on income. Concepts such as Q&Q methodology and SFD diagram were discussed, and how they could influence levels of pathogens at sampling locations.

Then we explained all other mapping data and discussed how it could affect levels of pathogens detected at sampling locations. This included: Socioeconomic group, population, waterways, sewer network, watershed delineation, safe management of excreta, public school, public toilets, market, health facilities.

Each sampling location was discussed in detail, including what experts knew about the area. Then the expected pathogen level on a scale of 1-5 specifically for the Kampala city context was discussed and agreed upon by the four experts.

- **5** indicates the highest pathogen level (“dangerous” was ranked in addition to 5 for the worst expected locations)
- **1** indicates the lowest (0 is none expected, lowest number in this study is 1)

At the end of the discussion, we reviewed together all 22 locations again and verified whether the original rankings were still held. 2-3 were adjusted at this time.

Following the Group discussion, we field validated the sites together. Sites 9, 1, 2, 3, 10, 4, 5, 11 were visited on 12 March 2025. The remaining sites are on 13 March 2025.

**Sampling Point 1 (0°21'02.6"N 32°33'38.7"E), Kawempe Division, Bwaise**

Expected pathogen level: **5**

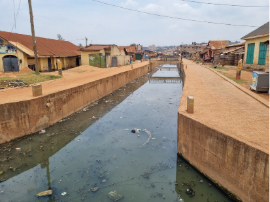

**Group discussion:** There is a major channel in this location that always floods, there is poor solid waste management, workers are required to clean the drains on a daily basis. Lots of flying toilets end up in this channel. This is a major informal settlement area of Kampala and is difficult for trucks to access for emptying.

**Field validation**: This was a slower moving channel right before a T entrance to the main channel. As they said, there are less toilets right on this channel now, as there was a World Bank project to remove them. Now it looks more like a sidewalk on the side of the channel. This area floods during the rainy season.

**Sampling Point 2 (0°21'54.6"N 32°34'24.8"E), Kawempe Division, Kanyanya**

Expected pathogen level: **3**

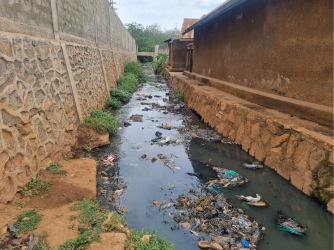

**Group discussion:** To the east of the sampling location is a high-income area, to the west are informal settlements. This received a “3” because of the mix of development types. The high-income area does not flood; trucks cannot access the informal settlements.

**Sampling Point 3 (0°21'53.6"N 32°34'39.1"E), Kawempe Division, Kyebando**

Expected pathogen level: **3**

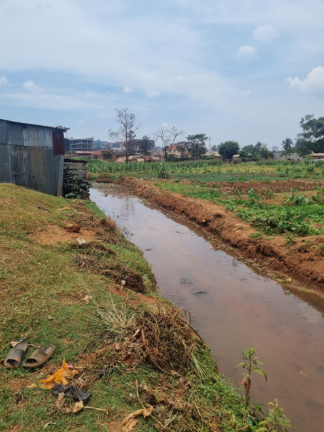

**Group discussion:** This is a hilly area; there are few informal settlements, and in these informal settlements there are more single homes / permanent structures. Although it is a informal settlement, it gets a 3 because it is not in such a sorry state and trucks can access here for sampling.

**Field validation**: Higher income than informal settlements, less densly populated

**Sampling Point 4 (0°21'23.8"N 32°35'25.2"E), Kawempe/Nakawa Division, Kyebando**

Expected pathogen level: **2 > 3**

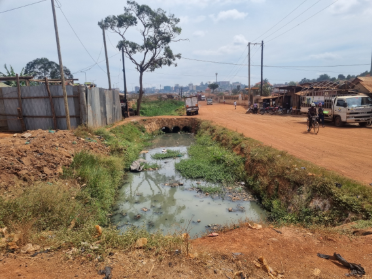

**Group discussion:** This was given a 2 because it is higher-end, and has been higher-end for a long-time. There are no hostels. Was planned to be ministers villas, and in the center some malls / trading centers / apartments

**Field validation**: Annet lowered this to a 3. It is more of an industrial area, some business parks, some apartments, did not seem interesting for sampling.

**Sampling Point 5 (0°22'59.8"N 32°36'29.5"E),**

**Kawempe/Nakawa Division,  Kyanja/Kulambiro**

Expected pathogen level: **2**

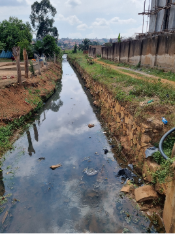

**Group discussion:** Rapidly growing, high end, lots of apartments. Kyanja has lots of apartments. Even though these apartments are connected to septic tanks, they are often just going directly to open drains, as the wastewater is way over capacity for the soak pits, it cannot fit in there.

**Field validation**: Not many houses, sparsely populated, on the boundary of Kampala

**Sampling Point 6 (0°20'14.5"N 32°32'04.0"E), Rubaga Division, Namugoona**

Expected pathogen level: **3**

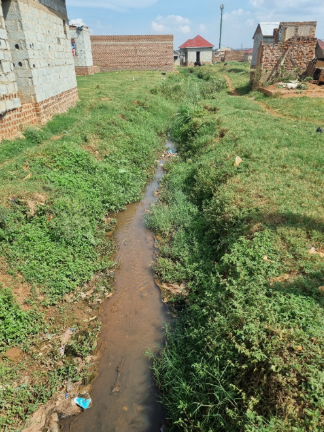

**Group discussion:** Lots of informal settlements. The group gave it a 3 because they are on a hill

**Field validation**: Could not get all the way there, but it looks accessible from the other side. Sampling point is at bottom, relatively steep going up on both sides. On the other side it looks like new houses are being built. It is a waterlogged area.

**Sampling Point 7 (0°20'24.7"N 32°32'53.0"E), Rubaga Division, Kawaala/Kasubi**

Expected pathogen level: **4**

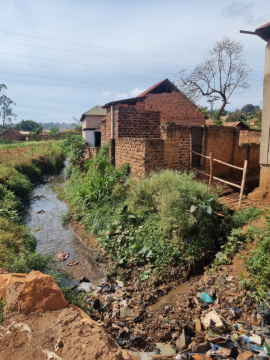

**Group discussion:** More informal settlements than SP 6, and it is lower elevation, and so they are in a worse position in the valley

**Field validation**: Road we went down is only road, not very accessible. Had to slightly move the SP for accessibility. SP is surrounded by gardens, a school, football field, upstream is denser informal settlements and mixed development. See also toilets draining to channel.

**Sampling Point 8 (0°20'34.0"N 32°33'27.7"E),**

**Rubaga/Central Division, Makerere Kikoni/Kawaala**

Expected pathogen level: **5 (“dangerous”)**

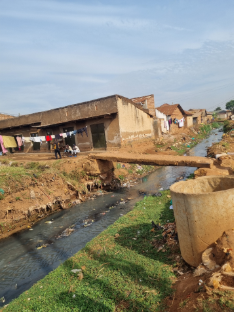

**Group discussion:** This was ranked as “dangerous”. There are lots of hostels which are releasing fresh fecal sludge into the environment, there is no capacity to contain this wastewater

**Field validation**: Fast-moving channel. There are many hostels near the west gate of Makerere.

**Sampling Point 9 (0°20'58.2"N 32°34'02.2"E),**

**Kawempe Division, Makerere Kavule/Mulago**

Expected pathogen level: **5 > 4**

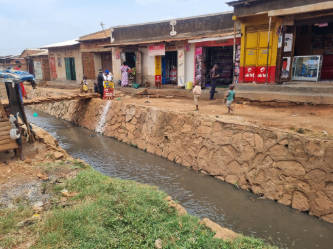

**Group discussion:** Katanga – this is a “real” informal settlements. Makerere is on the hill, slopes down to Katanga informal settlements, and then back up to Malaga hospital. This received a 5 because of Katanga, it is in a valley, and there are also many hostels around Makerere.

**Field validation**: There was confusion because in the Group discussion, it was thought we go to Katanga, which was a dangerous 5. However, where the Sampling point was is more Kalerwe, which is more like a 4 than a 5 because there are more separate houses with their own toilets.

**Sampling Point 10 (0°20'53.2"N 32°35'11.4"E), Nakawa/Central Division, Kamwokya**

Expected pathogen level: **5**

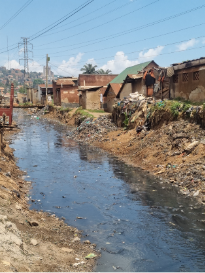

**Group discussion:** Used to be a wetland so there is a high-water table, encroached on lots of informal settlements. Above to the east is Kira Road / Acacia Mall / high end housing. In the lower income areas, the population is dense. The higher income areas are higher elevation. Even though it is a small area, the area is densely populated and “purely” informal settlements. Although part of this area is on the sewer (e.g., Makerere) there are also lots of informal settlements.

**Field validation**: High priority for sampling location. The channel divides the Passover, Central Division and the Mulimira, Nakawa Division

**Sampling Point 11 (0°20'20.3"N 32°37'13.8"E), Nakawa Div., Ntinda/Katalima**

Expected pathogen level: **3**

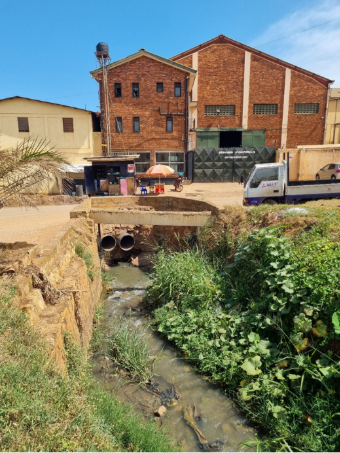

**Group discussion:** Mainly industrial, partly sewered. There are few residences, channel crosses straight through. NOTE: this was given a 3 by the group not because of pathogens, but because they were sure this is contaminated by industrial waste.

**Field validation**: VERY industrial, see the huge line of trucks, does not seem like an interesting sampling location.

**Sampling Point 12 (0°17'44.1"N 32°33'41.5"E), Rubaga Division, Rubiri**

Expected pathogen level: **2**

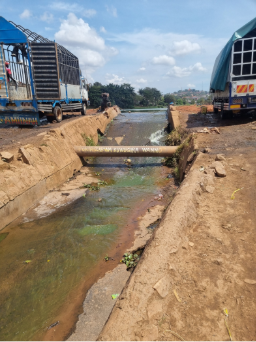

**Group discussion:** Above Masaka road is mostly residential, single units or only a few stories, there are key institutions like cathedrals .

**Field validation**: Outlet coming out of a man-made lake with small houses, apartments and hotels surrounding the lake. There are also informal settlements on the lake. The lake serves as a buffer. Downstream from the sampling point there are lots of auto repair type shops and the oil/grease/solvent pollution is quite strong, but it is downstream.

**Sampling Point 13 (0°18'37.5"N 32°34'28.8"E), Central/Rubaga Division, Kisenyi**

Expected pathogen level: **2**

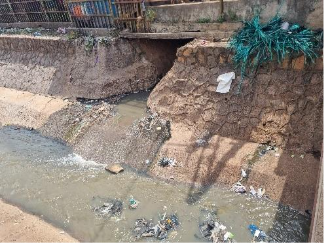

**Group discussion:** In this neighborhood is the protestant cathedral, but there are also lots of informal settlements, however they are higher-end and not dangerous. It is on a hill. The area just lower to this is sampling point 16.

**Field validation**: Right in the center of the busiest part of downtown at the Nakasero market next to the Old Taxi Park. 14 is the main channel, 13 is the side channel. 13 is covered in the market but could sample where it comes out into the channel.

**Sampling Point 14 (0°18'39.7"N 32°34'30.2"E), Central Division, Kisenyi**

Expected pathogen level: **4**

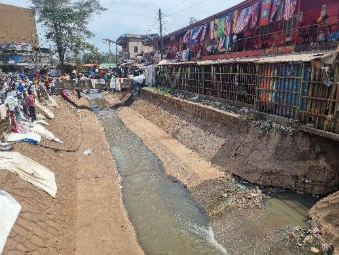

**Group discussion:** There are a lot of refugees here. The Gaddafi / National Mosque is located here (NOTE: can see from most parts of town). The majority of taxi parks are also located here, e.g. Kiseni taxi park, new taxi park.

**Field validation**: Quite crowded, should come early morning.

**Sampling Point 15 (0°19'03.0"N 32°35'38.7"E), Central Division, Nakasero/Kololo**

Expected pathogen level: **1**

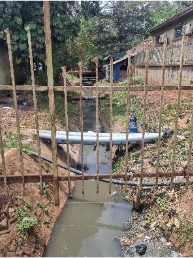

**Group discussion:** This is a sewered area; however, there are also informal areas that are off the sewer/they cannot afford to connect to the sewer. To the west of the sampling point is an informal area that drains to the channel.  Nakasero and Kololo would be ranked 1, but the problem is that the sewer line from the hospital and another from the hotel are not properly contained.  The State House is in this area, and the best hotels are all in this area. The problem with ranking this area is that it is a HUGE area.

**Field validation**: The Centennial Park is upstream of sampling point, here there are bars, shops, etc. Kololo is also upstream. The site is on a busy road / bridge / highway. There is a public toilet right there that looks very clean and well maintained

**Sampling Point 16 (0°17'29.7"N 32°33'44.6"E), Rubada Division, Nalukolongo/Ndeeba**

Expected pathogen level: **5**

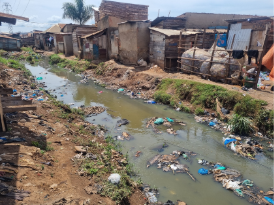

**Group discussion:** This is where there was a Cholera outbreak in 2017, very informal, a lot of solid waste issues, the channel is always clogged, there is also a market.

**Field validation**: The railway line separates Koboa and Ndeeba in Rubaga, the canal is on the Koboa side. The sampling point was further down, but it was inaccessible for driving (road fell away from railroad tracks), but this point would be OK. The is a “real” informal settlement, we obviously need to distinguish between low and very-low income.

**Sampling Point 17 (0°18'17.9"N 32°34'37.2"E),**

**Central/Makindye/Rubaga Division, Katwe/Kibuye**

Expected pathogen level: **4**

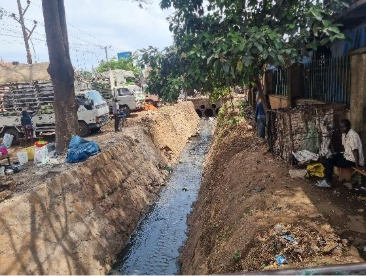

**Group discussion:** Although there are informal settlements, they are more established with property ownership and permanent buildings. In Kampala, if you have occupied the land for 12 years, you have rights to ownership. However, there is also the question of the major landowner, which is the Buganda Kingdom.

**Field validation**: Right at Usafi Market. Down from the main taxi park. It seemed relatively deep. Maybe it is a transitory population.

**Sampling Point 18 (0°18'31.3"N 32°35'22.4"E), Makindye Division, Kibuli**

Expected pathogen level: **3**

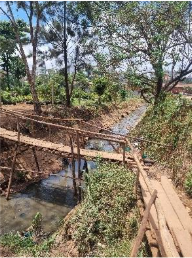

**Group discussion:** This is the first place historically where muslims settled in Kampala. There are lots of informal settlements.

**Field validation**: At this sampling point the road/bridge was under construction; the downstream side did not seem like a good sampling location due to trash and accessibility. However, the upstream side was accessible. At this location there is a police school. There are garden plants along the bank, and it is adjacent to the school playground. It is not very densely populated but maybe upstream populations. There are informal settlements, but we did not see whether it is on the canal.

**Sampling Point 19 (0°16'42.5"N 32°33'16.8"E), Rubaga Division, Wankulukuku**

Expected pathogen level: **5**

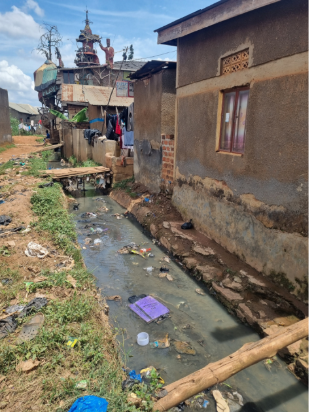

**Group discussion:** Wankulukuku is where the Cholera outbreak started in 2017. This is very much an informal settlement area; it is a low-lying area with high groundwater and poor Solid waste management.

**Field validation**: Near the stadium. The water in the channel is dirty.

**Sampling Point 20 (0°16'42.3"N 32°34'37.1"E),**

**Rubaga/Makindye Division, Najjanankumbi/Makindye**

Expected pathogen level: **3**

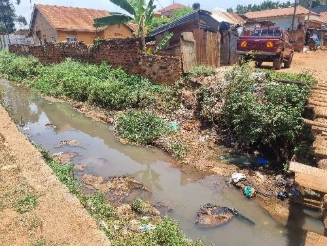

**Group discussion:** Makindye, half Najanankumbi and half Makindye. Salama Road and Busabala Road. This is on the edge of Kampala city, there are lots of informal settlements, but not as dense and on a hill.

**Field validation**: Quite steep downhill to the point. The car couldn’t get in. Had to walk 2 minutes.  There were also newer/under construction houses on the other side.

**Sampling Point 21 (0°17'19.6"N 32°35'40.5"E) Makindye Division, Nsambya**

Expected pathogen level: **4.5**

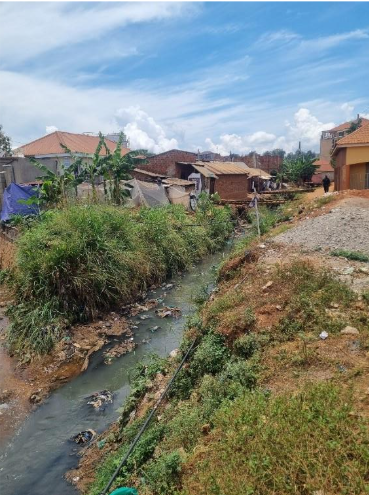

**Group discussion:** There are many toilets along the edge of the channel.

**Field validation**: SP 21 and 22 are right next to each other. 21 is the main channel and is more middle-income. 22 is the side channel and is more low-income

**Sampling Point 22 (0°17'18.0"N 32°35'41.8"E), Makindye Div., Nasambya**

Expected pathogen level: **3.5**

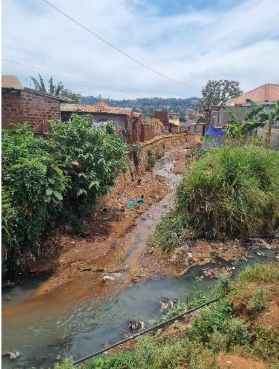

**Group discussion:** This is a mix. Lots of high-income people, apartments, and informal settlements.

**Group discussion on April 1, 2025**

**Experts at Kampala Capital City Authority (KCCA), Makerere University, and Eawag**

Ten sampling points were discussed again after sharing the field validation and pictures.

1. Reconfirmed/adjusted the consensus expected pathogen level (**1-5**).
2. Based on the field validation, a new socioeconomic category was introduced, including **very-low income**, **low-income**, and **middle-income**

The table below summarizes the pathogen level and socioeconomic status of 10 sampling points (#1, 8, 9, 10, 13, 14, 16, 19, 21, 22). The detailed discussion on each point is followed.

| Sampling points | Division | Parish/village | Expected pathogen level | Socioeconomic status |
| --- | --- | --- | --- | --- |
| 1 | Kawempe | Bwaise/Nakamiru | 5 | Very low-income |
| 8 | Rubaga/Central | Makerere Kikoni | 5 | Middle-income |
| 9 | Kawempe | Makerere Kavule | 5 | Low-income |
| 10 | Nawaka/Central | Kamwokya | 5 | Very low-income |
| 13 | Central/Rubaga | Kisenyi | 3.5 | Commercial |
| 14 |  |  | 5 |  |
| 16 | Rubaga | Ndeeba | 5 | Very low-income |
| 19 | Rubaga | Wankulukuku | 5 | Very low-income |
| 21 | Makindye | Nsambya Gogonya | 4.5 | Middle-income |
| 22 |  | Nsambya Lukuli | 3.5 | Middle-income |

 #1 – Bwaise, Nakamiru

It used to be quite a flood-prone area before the World Bank constructed the channel. There was a discussion about whether it is still often flooded a lot. But, it was agreed that the pathogen level and socioeconomic status are “**5**” and “**very low-income**”

 #8 – Makerere Kikoni

The unique environment here, where many hostels are located. The expected pathogen level is “**5**” due to poor sanitation. Jude said it could be “4”.  But, socioeconomic status is at a more “**middle-income level**”.

 #9 – Makerere Kavule

Upstream (mini-catchment) is *Katanga*, where sanitation is terrible. The sampling point is located at *Kalerwe*. Even though the field validation suggested “4”, all agreed that it should still be “**5**”. Socioeconomic status was agreed to be “very low-income” at the first discussion but after comparing other “very low-income” points, it was decided as “**low-income**”.

#10 – Kamwokya

Kamwokya, fecal matter is everywhere. Pure informal settlements, easily agreed on pathogen level of “**5**” and “**very low-income**” area.

 #13 (side) and #14 (main) – Nakivubo channel, market

This is quite a distinct area compared to other sampling points. Commercial and transitory areas but also numerous homeless people (highest percentage in Kampala) reside. The socioeconomic status cannot be decided with the existing category. Just leave as “**Commercial**”.

In the previous discussion, it was said that the Gaddafi / National Mosque is located here. But, it was corrected that this Mosque is located downstream. So, sampling here doesn’t reflect that area.

Micro-catchment of #13 includes the new park, a stadium, and Rubaga hospital. Some drains go directly to the lake. The expected pathogen level is “**3.5**”

Micro-catchment of #14, lots of homeless. The expected pathogen level is “**5**”.

 #16 – Ndeeba

Pure informal settlements, easily agreed on pathogen level of “**5**” and “**very low-income**” area.

 #19 – Wankulukuku

Pure informal settlements, easily agreed on pathogen level of “**5**” and “**very low-income**” area.

 #21 and #22 – Nsambya

#21 Gogonya, apartment nearby, but still frequent discharging to the drain is expected

Pathogen level “**4.5**”, “**middle-income level**”

#22 Lukuli, church in high-end area

Pathogen level “**3.5**”, “**middle-income level**”

NOTE: There was a concern that if we didn’t address high-income level in the sampling points, it could induce incorrect perceptions in people. Thus, it should be noted that sampling points were selected with the intention to monitor vulnerable communities in Kampala. Still, there was a discussion about whether some high-income areas should be included for comparison.

Also, since no Nakawa division was covered by 10 sampling points, the information in this division will be lacking. It should be noted that this should be considered in planning the next campaign. It is noted that all socioeconomic category here is only applied within Kampala, not being compared to other countries.
